# The Effects of Gender Affirming Hormone Treatment on Transgender Men’s Musculoskeletal Health: A Systematic Review and Meta-Analysis

**DOI:** 10.64898/2026.03.31.26349844

**Authors:** Ke Hu, Andrew Brown, Stephanie Montagner-Moraes, Jatinder Singh, Laura Charlton, James Barrett, Blair R. Hamilton

## Abstract

**Background:** A previous meta-analysis by Singh-Ospina et al. (2017) suggested that Gender affirming hormone treatment (GAHT) does not change transgender men’s bone mineral density (BMD) at any clinically relevant site; emerging studies and advances in synthesis methods necessitate an updated evaluation. The primary aim was to update the bone measures of Singh-Ospina et al. (2017), with the secondary aim to expand measures to how GAHT affects musculoskeletal health.

**Methods:** A systematic review with meta-analysis was conducted using studies published in English up to 31 July 2024, identified through three electronic databases (PubMed, Embase, SportDiscus), and final cross-referencing in summer 2025. Primary outcomes were longitudinal changes in femoral neck (FN), lumbar spine (LS), and total hip (TH) bone mineral density (BMD). Secondary outcomes included body composition and muscle strength. Standardised effect sizes (Hedges’ *g*) were pooled using the inverse heterogeneity (IVhet) model.

**Results:** GAHT (4 ± 5 years) was not associated with significant longitudinal changes in FN, LS, or TH BMD. In contrast, substantial anabolic effects were observed, including increases in BMI (*g* = 0.13), body mass (*g* = 0.18), fat-free mass (*g* = 0.59), and muscle strength (*g* = 0.86). Heterogeneity was high for muscle strength, FN and TH BMD, limiting confidence in pooled estimates. Conversely, changes in LS BMD, BMI, body mass and fat-free mass demonstrated low heterogeneity and greater consistency across studies.

**Conclusion:** Masculinising GAHT does not negatively affect clinically relevant BMD sites while reliably increasing lean mass and muscle strength; however, the evidence base remains methodologically weak and highly variable, particularly for FN and TH. The need for continued clinical monitoring of bone health and muscle function, alongside high-quality longitudinal research incorporating advanced imaging modalities such as HR-pQCT is emphasised. Strengthening the evidence base will be essential for clarifying long-term skeletal trajectories as transgender men age.

**PROSPERO registration:** CRD42024573102

## Introduction

Transgender men may experience psychological distress related to a mismatch between their assigned gender at birth and their experienced gender, a condition sometimes referred to in medical contexts as gender dysphoria (Jones et al., 2017; Kuyper & Wijsen, 2014). Approximately 3% of the global population, equating to around 241 million people (Statista, 2023) identify as transgender. Many of these individuals undergo medical interventions, including gender-affirming hormone treatment (GAHT) and gender-affirming surgery, to align their physical characteristics with their gender identity (Coleman et al., 2012; Wylie et al., 2014). The demand for transgender and gender diverse health services has notably increased globally in recent years (Aitken et al., 2015; Arcelus et al., 2015) reflecting increased recognition of the healthcare needs of transgender and gender diverse identities.

For transgender men (individuals assigned female at birth who identify as male), GAHT usually involves testosterone administration, which has been shown to reduce distress and increase the quality of life in transgender men (Foster Skewis et al., 2021; Murad et al., 2010; White Hughto & Reisner, 2016). Androgens like testosterone are essential for maintaining skeletal homeostasis by stimulating osteoblast activity, suppressing osteoclast-mediated bone resorption, and supporting muscle mass, thereby enhancing mechanical loading on bone (Raggatt & Partridge, 2010; Xu et al., 2022). In contrast, oestrogen plays a pivotal role in preserving cortical bone (which is 80% of the bone mass) by regulating bone remodelling and limiting excessive bone turnover (Aguirre et al., 2007; Cole et al., 2008; Ji & Yu, 2015; Kasperk et al., 1997; Mohamad et al., 2016). In transgender men undergoing GAHT, exogenous testosterone alters the endogenous sex steroid environment, including changes in circulating oestrogen levels via aromatisation. These shifts may differentially affect trabecular and cortical bone compartments, potentially influencing bone density, geometry, and strength over time. Consequently, long-term alterations in sex steroid exposure may have clinically relevant implications for musculoskeletal health, particularly with ageing and prolonged duration of hormone therapy. The coordinated regulation of androgens and oestrogens is critical for preserving skeletal integrity; therefore, alterations of sex steroids may exert complex effects on bone metabolism. Consequently, the implications of sex steroids on bone health must be carefully considered in transgender men. However, the effects of GAHT on musculoskeletal health are less understood, highlighting the need for further research and regular monitoring of bone density and hormonal profiles.

A previous meta-analysis by Singh-Ospina et al. (2017) examined the effects of GAHT on bone health in transgender men, The authors found no statistically significant difference in lumbar spine (LS), femoral neck (FN), and total hip (TH) bone mineral density (BMD) following short-term hormone therapy at 24 months. The authors concluded that in transgender men, GAHT was not associated with an increase in BMD at the LS (0.00 g/cm^2^), FN (0.02g/cm^2^), or TH (-0.01 g/cm^2^). While the results reported by Singh-Ospina et al. are noteworthy, they were limited to 13 trials, observational studies, and case series published up to April 2015, lacked an assessment of BMD using quantitative computed tomography (QCT), and did not assess muscle health.

Since Singh-Ospina et al. (2017), additional studies have been published (Bobba et al., 2023; T. M. Fighera et al., 2018; Yun et al., 2021) and more robust methods for the undertaking and interpretation of meta-analytic results have been developed (Doi et al., 2015; Doi et al., 2017; Furuya-Kanamori et al., 2018; VanderWeele & Ding, 2017).Cheung et al (Cheung et al., 2024) reported an 19% increase in muscle mass within three years and 18% increase in strength in one year, and a comparable physical performance level of cisgender men by the third year of testosterone therapy; however, without employing a meta-analytic approach and primarily focusing on sports performance rather than musculoskeletal health-related. Finally, using previously developed guidelines for when to update a systematic review, it was decided that an updated review on this topic was needed (Garner et al., 2016).

Thus, given 1) the updated preferred reporting items for systematic reviews and meta-analyses (PRISMA) statement (Sarkis-Onofre et al., 2021) 2) the potential effects of GAHT on transgender men’s musculoskeletal health outside of BMD, 3) the lack of recent meta-analytic work in this area, and 4) the use of more robust methods for conducting meta-analytic research (Doi et al., 2015; Doi et al., 2017; Furuya-Kanamori et al., 2018; VanderWeele & Ding, 2017) and 5) decision tree analysis of when to update a systematic review (Garner et al., 2016) We aimed to update and expand on the systematic review and meta-analysis by Singh-Ospina et al. (Singh-Ospina et al., 2017), whereby we examined the effects of GAHT on transgender men’s musculoskeletal health.

## Materials and Methods

### Study Eligibility Criteria

As this meta-analysis aims to update the Singh-Ospina et al. (Singh-Ospina et al., 2017), the same *a priori* inclusion criteria were employed (**Table 1**), with additional studies identified from 07/04/2015. Studies that do not meet the criteria outlined in Table 1 were excluded from the analysis. This meta-analysis adhered to the PRISMA guidelines (Moher et al., 2009). The protocol was preregistered in PROSPERO (trial registration number: CRD42024573102 (Brown et al., 2024b)).

**Table 1.**
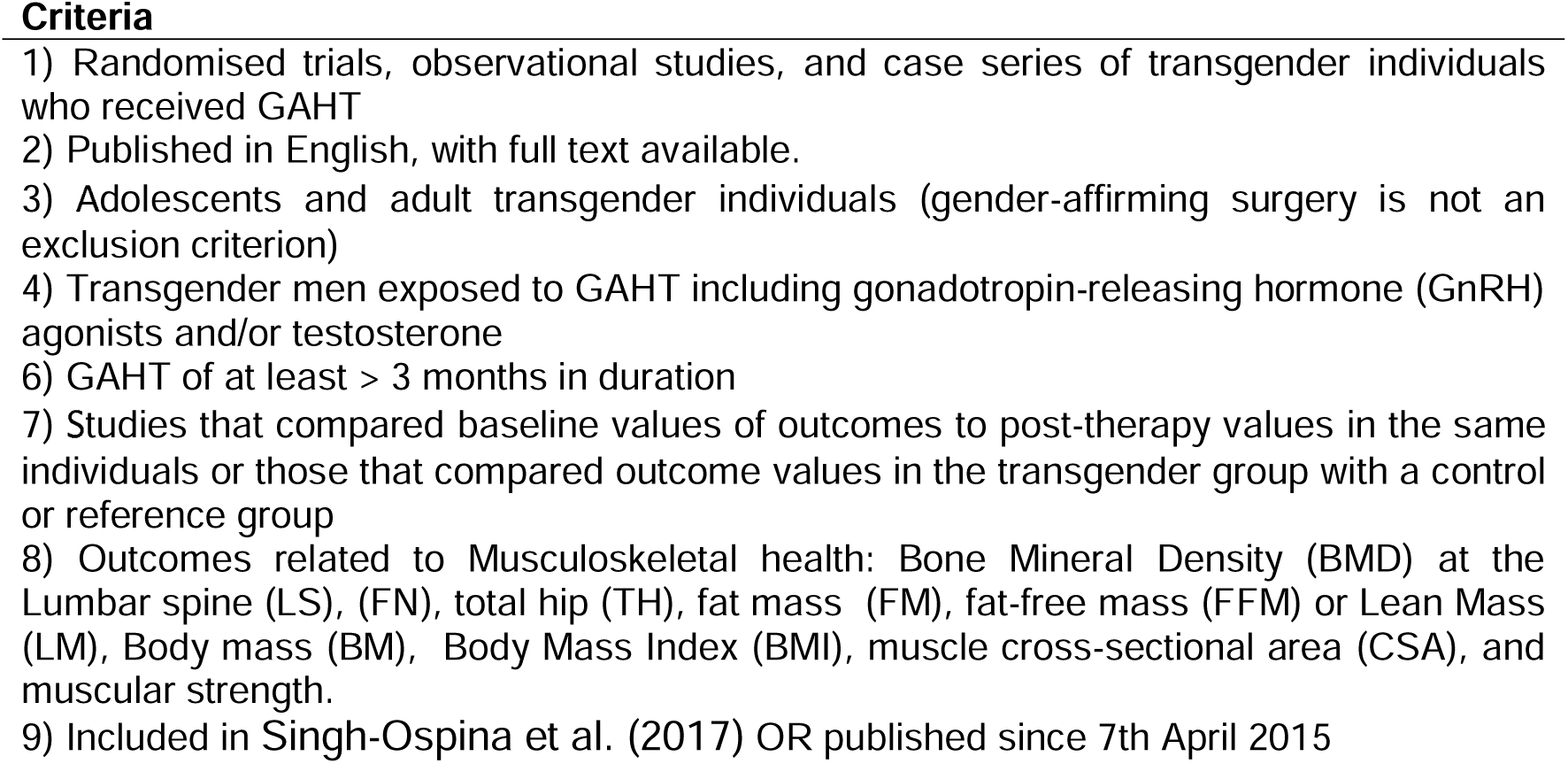
Study Eligibility Criteria.

### Data Sources

Studies published up to 31^st^ July 2024 were retrieved from three electronic sources (PubMed, Embase, SportDiscus). Keywords relevant to all searches included “transgender,” “bone,” and “muscle.” A final database search was conducted on 1^st^ December 2024. Based on PRISMA guidelines (Moher et al., 2009), an example of the search strategy is supplied in the supplementary material (A. Brown et al., 2025). The last author (BRH) conducted all electronic database searches. In addition to electronic database searches, cross-referencing from retrieved studies was also conducted.

### Study Records and Selection

All studies were imported into EndNote (EndNote 20.6, Clarivate Analytics, USA), and the last author (BRH) removed duplicates electronically and manually. A copy of the reference database was then provided to the second author (AB) and the third (SMM) for dual screening. Both authors (AB and SMM) screened studies independently. The screeners were not blinded to journal titles or the study authors/affiliations. Reasons for exclusion were coded based on one or more of the following: 1) inappropriate population, 2) inappropriate intervention, 3) inappropriate comparison(s), 4) inappropriate outcome(s), 5) inappropriate study design or 6) other. On completion, the screeners met to discuss their selections and reconcile any discrepancies by consensus. If an agreement cannot be achieved, the last author (BRH) provided a recommendation. The agreement rate, before the reconciliation of any discrepancies, was calculated using Cohen’s κ statistic (Cohen, 1968). The precision of searches was calculated as the number of studies included divided by the number of studies screened (less duplicates) (Lee et al., 2012). The number needed to screen (NNS) was calculated by taking the reciprocal of the precision (Lee et al., 2012).

### Data Abstraction

Before data abstraction, an electronic codebook was developed by the last author (BRH) and provided to KU/AB/SMM. The extracted data were coded based on the following major categories: 1) study characteristics (e.g., author, journal, year, etc.), 2) participant characteristics (e.g., age, height, mass, etc.), 3) intervention details (e.g., type, length, frequency, etc.), and 4) outcome characteristics (e.g., sample sizes, baseline/post-GAHT means and SD, etc.). A copy of the electronic codebook is available in the supplementary material (A. Brown et al., 2025).

The first (KH), second (AB) and third (SMM) authors extracted all data independently of one another before meeting to resolve any discrepancies by consensus. If an agreement was not achieved, the last author (BRH) provided a recommendation. Before this, the overall agreement rate was assessed by Cohen’s κ statistic (Cohen, 1968).

### Outcome Measures

*A priori* primary outcome measures were changes in bone health parameters such as Total Hip (TH), Femoral Neck (FN) and Lumbar Spine (LS) bone mineral density (BMD) measured by Dual-Energy X-Ray Absorptiometry (DXA), Dual-Photon Absorptiometry (DPA), or Quantitative Computed Tomography (QCT). Secondary, *a priori* outcomes included changes in body mass (BM), body mass index (BMI), Lean body mass (LM) or fat-free mass (FFM), fat mass (FM), muscle cross-sectional area (CSA) and muscular strength. Obtaining missing data was attempted for all primary and secondary outcome measures if assessed by a study and the data provided proved inadequate to calculate an effect size. The last author (BRH) contacted the study’s corresponding author three times via email, with one week between each communication. These communications were tracked (e.g., dates, responses, success rates, etc.) to establish the success rate of this process.

### Risk of Bias Assessment

The risk of bias for each study was assessed using the recently revised Cochrane Risk of Bias instrument for non-randomised longitudinal studies of interventions (ROBINS-I) and JBI Critical Appraisal Tools for cross-sectional studies (J. A. Sterne et al., 2016). Using one or more signalling questions, the ROBINS-I instrument assessed the risk of bias in seven distinct domains: (1) bias arising from confounding, (2) bias in participant selection (3) bias in classification of interventions (4) bias due to deviations from intended interventions, (5) bias due to missing data, (6) bias in the measurement of outcomes and (7) bias in the selection of the reported result. Based on signalling questions, each domain was assessed as either ‘low risk,’ ‘moderate risk,’ ‘serious risk,’ or ’critical risk.’ Based on responses to each domain, the overall risk of bias for each study was assessed as either ‘low risk,’ ‘moderate risk,’ ‘serious risk,’ or ’critical risk.’ JBI checklist includes eight items covering sampling, measurement validity, confounding, and statistical analysis. Each item was rated as ‘Yes’, ‘No’, ‘Unclear’, or ‘Not applicable’. An overall risk of bias judgment was then made for each study based on these domains. We chose to use this risk of bias instrument over the various study quality instruments, including those focused on intervention studies (Maher et al., 2003; Smart et al., 2015) given the difficulty of the latter in differentiating between the quality of reporting and the quality of the conduct of a study (J. A. C. Sterne et al., 2019).

No studies were excluded from the analysis based on the risk of bias assessment (Ahn & Becker, 2011). Our decision not to exclude on risk of bias was guided by the recognition that the field of transgender musculoskeletal health remains in its early stages, with a relatively limited number of available studies, and the inability to blind participants to their GAHT. Excluding all studies with a serious or critical risk of bias would have severely restricted the scope of the analysis and potentially overlooked emerging patterns in the literature. To address this limitation transparently, we conducted sensitivity analyses where possible and explicitly reported the risk of bias assessments alongside study findings. This approach allowed us to highlight the strength of evidence while also acknowledging its limitations.

### Statistical Analysis

#### Calculation of effect sizes

The *a priori* primary and secondary outcomes for this meta-analysis were calculated using the Hedges standardised mean difference effect size, *g*, adjusted for small sample sizes (Hedges & Olkin, 2014). The *g* for each group was calculated as the mean of the baseline measure or the control/reference group minus the mean of the GAHT intervention group, divided by the pooled and weighted standard deviation. If this information is unavailable, *g* was calculated using procedures described by Follmann et al. (1992). For studies reporting multiple post-intervention time points, *g* was calculated using the baseline and final time points to report the longer-term effects of GAHT on musculoskeletal health.

#### Effect size pooling

Results were pooled using the inverse heterogeneity (IVhet) model (Doi et al., 2015), a model which is more robust than the Der Simonian–Laird random effects method employed by Singh-Ospina (Singh-Ospina et al., 2017). Two-tailed z-scores with alpha <0.05 and non-overlapping 95% confidence intervals will be considered statistically significant. For the longitudinal analysis, outcomes were compared at baseline and at the furthest available time point from baseline within the eligible study. This approach allowed us to assess changes over time within the same individuals. In the cross-sectional analysis, the eligible study must have included a cisgender female comparison group alongside the transgender male group. This comparison was used to evaluate differences between the groups at a single time point, providing a snapshot of the relative outcomes between the two populations.

#### Heterogeneity and Inconsistency

For each pooled outcome, heterogeneity was assessed using Q (Higgins et al., 2003), with an alpha level of <0.10 representing statistically significant heterogeneity. Inconsistency was assessed using *I^2^*, an extension of Q. For this meta-analysis, inconsistency was categorised as very low (<25%), low (25-50%), moderate (50-75%) or large (>75%) (Higgins et al., 2003). Absolute between-study heterogeneity was assessed using tau squared (^2^). In addition, influence analysis was conducted by removing each study from our analysis once to examine the effect of that study on the overall findings. Given the expected small sample size, no subgroup or meta-regression analysis was planned a *priori*.

#### Meta-biases

Small-study effects (publication bias, etc.) were assessed qualitatively using the Doi plot and quantitatively using the Luis Furuya-Kanamori index (LFK index) (Furuya-Kanamori et al., 2018; Furuya-Kanamori et al., 2020). The Doi plot has been suggested to be more intuitive than the funnel plot, and the LFK index is more robust than the commonly used Egger’s regression-intercept test (Furuya-Kanamori et al., 2018; Furuya-Kanamori et al., 2020). LFK values within ± 1, greater than ± 1 but within ± 2, and greater than ± 2 were considered to represent no, minor, and major asymmetry (Furuya-Kanamori et al., 2018).

#### Strength of evidence

The strength of findings for each outcome was assessed using the most recent version of the Grading of Recommendations Assessment, Development and Evaluation (GRADE) meta-analysis tool (Schünemann et al., 2013). Quality of evidence was assessed across the domains of risk of bias, consistency, directness, precision, and publication bias. Quality was judged as high (further research is very unlikely to change our confidence in the estimate of effect), moderate (further research is likely to have an important impact on our confidence in the estimate of effect and may change the estimate), low (further research is very likely to have an important impact on our confidence in the estimate of effect and is likely to change the estimate), or very low (very uncertain about the estimate of effect) (Schünemann et al., 2013)

#### Software used for analysis

All data were analysed using MetaXL (version 5.3, Epigear International Pty Ltd). All data is available as supplementary material (Brown et al., 2024a).

## Results

### Study Characteristics

A flow diagram that depicts the search process for study selection is shown in **Figure 1**. A full list of studies is available in the **Supplementary file TransMA_BH_Combined (Brown et al., 2024a).** After identifying 1157 citations, removing 32 studies due to publication before 7th April 2015 and 42 duplicates both electronically and manually, 1083 studies were screened. Of the 1083 studies reviewed, thirty-eight additional studies were included in this meta-analysis, bringing the total number of studies to 55. The results of transgender women were published in our sister meta-analysis to provide the reader greater clarity in results and discussion (Andrew Brown et al., 2025). A post-hoc decision was made to exclude four studies on transgender adolescents under 18 years old (Klaver et al., 2018; Tack et al., 2018; M. A. van der Loos et al., 2021; Vlot et al., 2017), due to the confounding effects of puberty on musculoskeletal health. Therefore, twenty-eight studies, 8 from Singh-Ospina (Singh-Ospina et al., 2017). and 20 additional studies were included in this analysis, where 2361 participants were transgender men, and 85 participants were comparator cisgender women.

**Figure 1.**
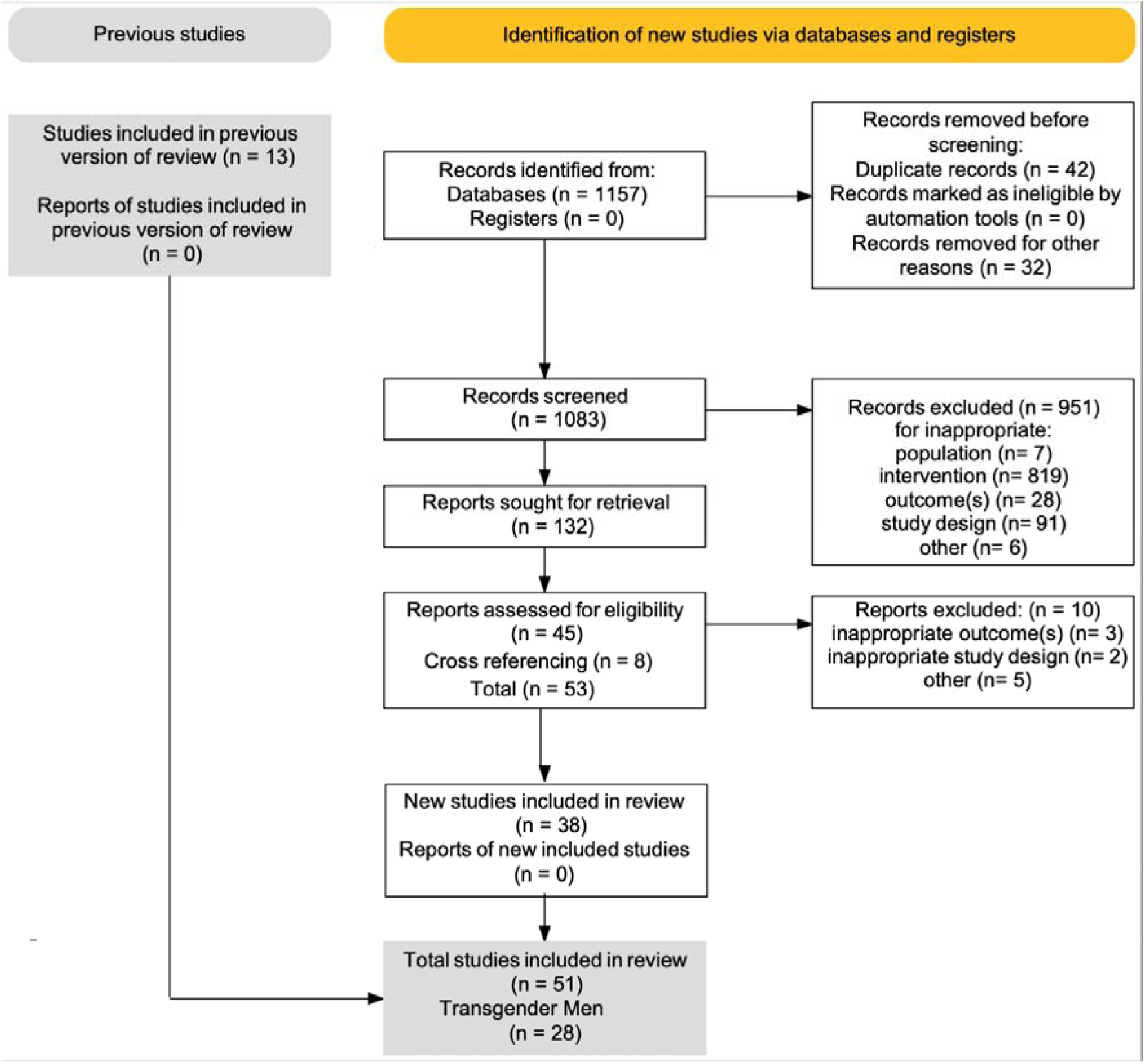
PRISMA Flow Chart.

The agreement rate between assessors for inclusion was 0.92 before reconciling any discrepancies, with all discrepancies resolved by consensus between the first, second and third authors. The major reasons for exclusion were inappropriate population (0.8%), inappropriate intervention (86.2%), inappropriate comparison (0.7%), inappropriate outcome(s) (3.0%), inappropriate study design (9.6%) and other (0.1%). The precision of searches was 12% while the number needed to screen was eight. A list of excluded studies can be found in Supplementary File 1, with a general description of the studies included found in **Table 2**.

**Table 2.**
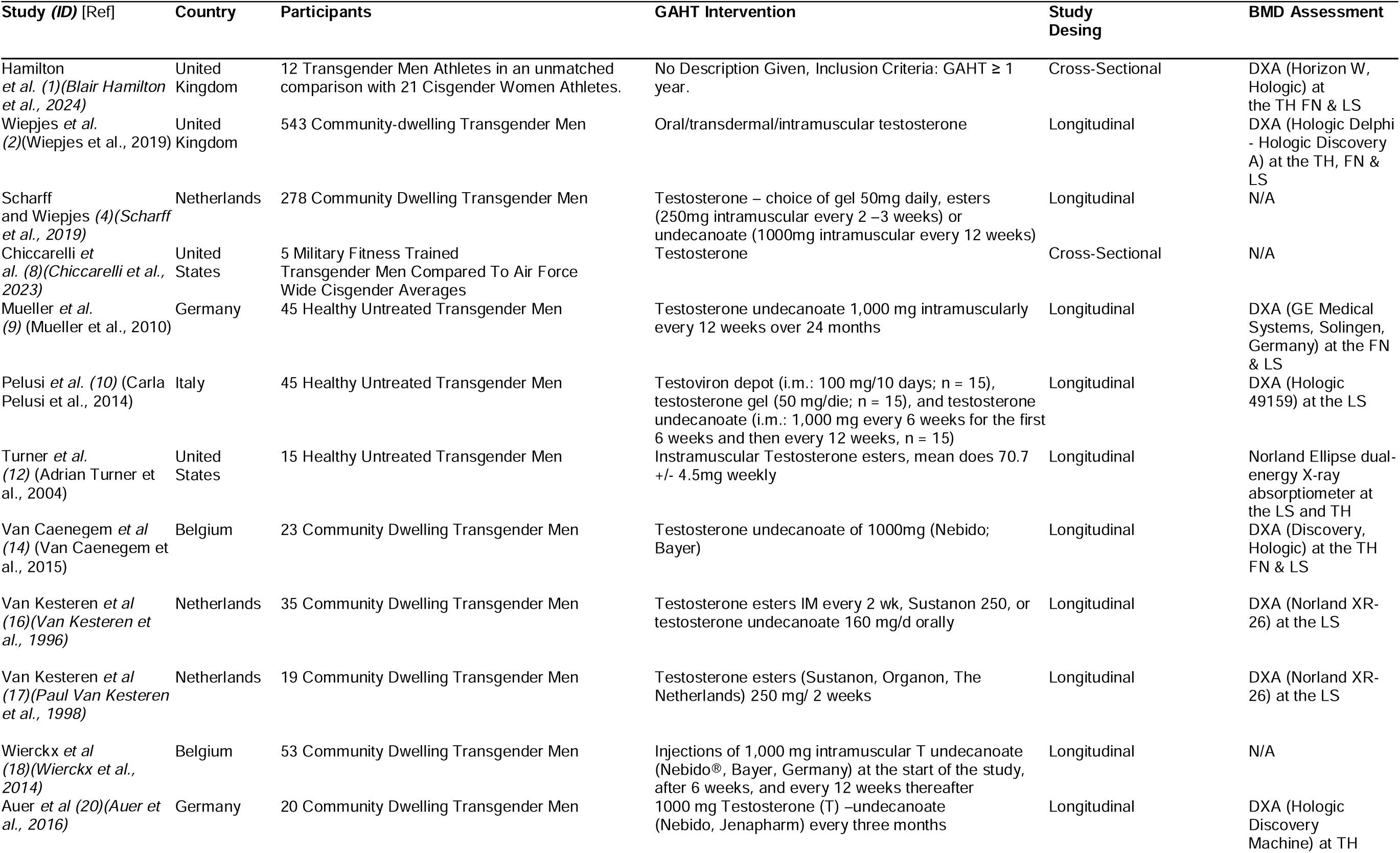
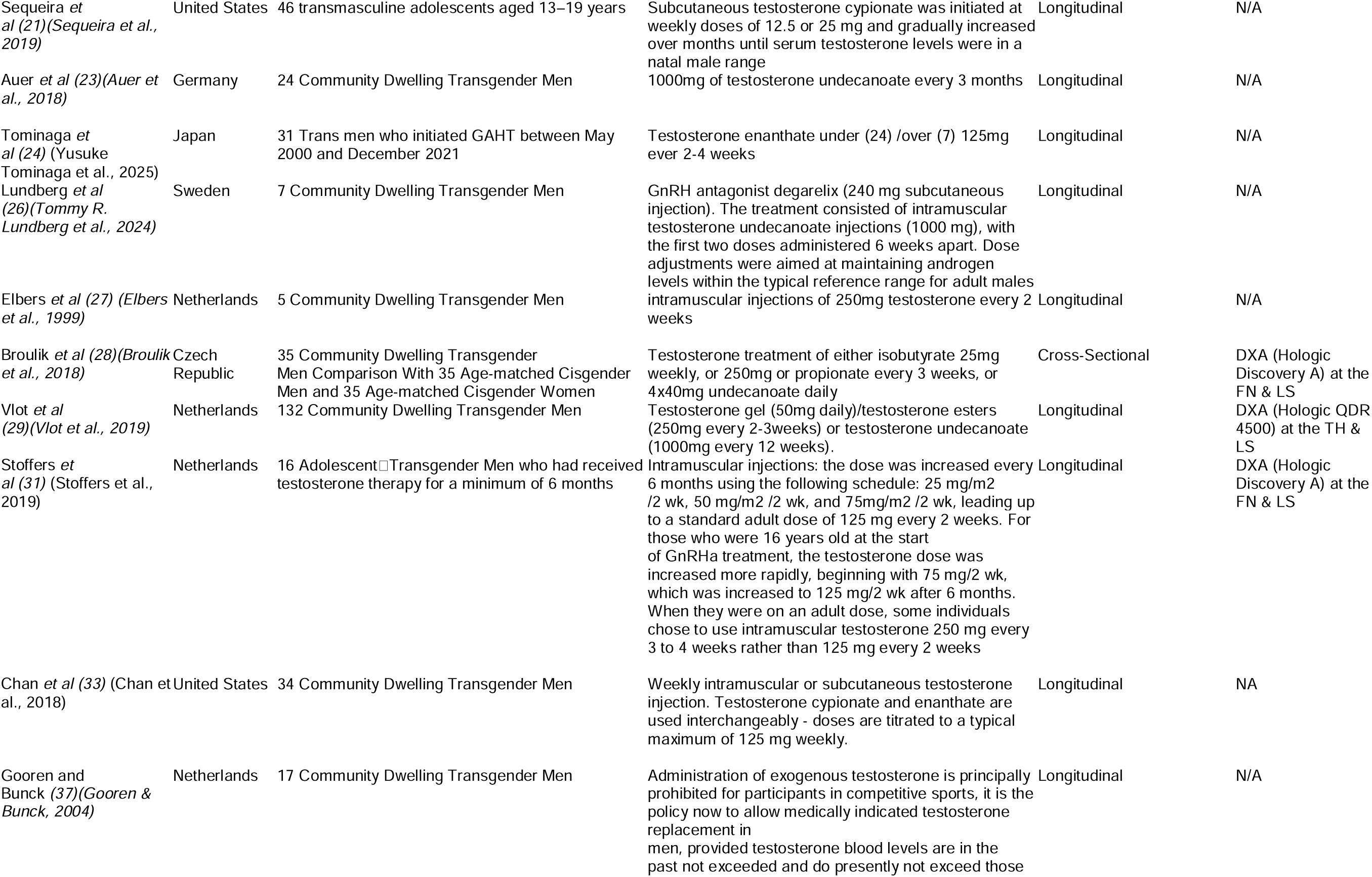
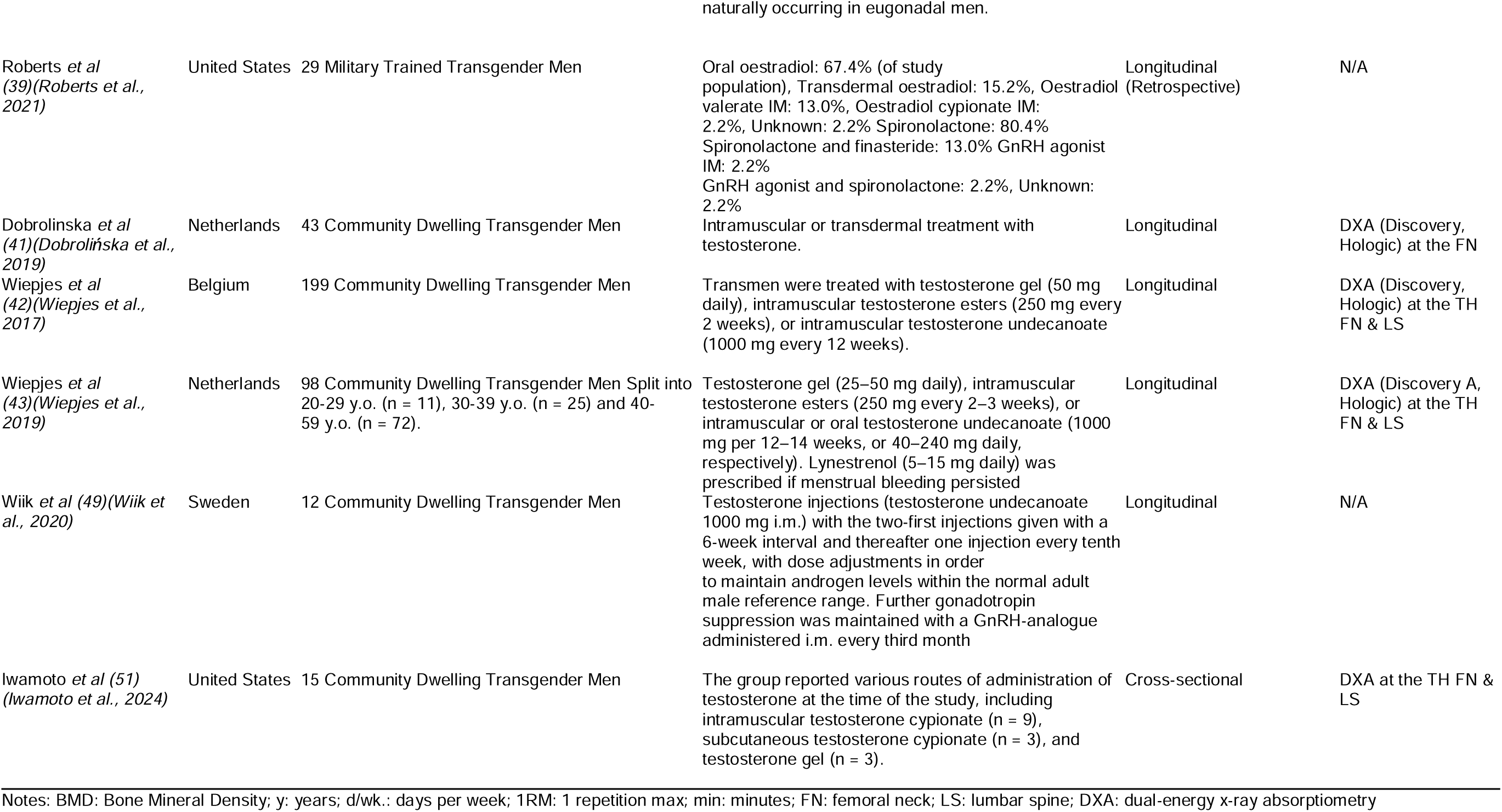
General characteristics of the studies included (*n* = 28)

Of the twenty-eight recent studies included, all were published in peer-reviewed English-language journals, starting in 2016 and ending in 2024. Two studies published before 2016 were included as they were identified during cross-referencing to include outcomes for muscle and body composition that were in the inclusion criteria and not part of the original meta-analysis by Singh-Ospina (Singh-Ospina et al., 2017). The studies were conducted in a variety of countries; nine in the Netherlands, six in the United States, three in Belgium and Germany, two in Sweden and the United Kingdom, and one each in Italy, Japan and the Czech Republic. The maximum number of Transgender men for which musculoskeletal health assessment was available ranged from 5 to 543 in the intervention groups (mean ± SD= 55 ± 99, [median = 21]) and from 8 to 50 in the comparison groups (mean ± SD = 26 ± 11, [median =24]). Two studies provided sample size estimates, while the rest did not. The agreement rate on data extraction was 1.0, with no discrepancies between the authors. Two studies (Bretherton et al., 2022; M. van der Loos et al., 2023) did not include all data within the manuscript and after three emails to the corresponding author, there was no response.

### Participant Characteristics

A description of the baseline characteristics of participants can be seen in **Tables 2** and **3**. Reported dropout rates ranged from 0 to 96.2% in the transgender men groups (mean ± SD = 31.8± 36.2%, median 6.6%), and there were 0% dropouts in the comparison groups. Twenty per cent of cross-sectional comparisons of transgender men to cisgender women were matched (age and BMI), compared to 80% unmatched.

**Table 3:** Baseline Characteristics of Participants.

| Variable | Transgender Men |  |  |  |  | Cisgender Women Comparators |  |  |  |  |
| --- | --- | --- | --- | --- | --- | --- | --- | --- | --- | --- |
|  | Groups (#) | Participants (#) | Mean ± SD | Mdn | Range | Groups (#) | Participants (#) | Mean ± SD | Mdn | Range |
| Age (y) | 29 | 2361 | 28 ± 6 | 27 | 16 - 47 | 2 | 47 | 35 ± 8 | 30 | 30 – 49 |
| BMI (kg/m<sup>2</sup>) | 17 | 975 | 23.4 ± 1.3 | 23.5 | 21.5 – 26.2 | 2 | 47 | 28.2 ± 4.9 | 27.5 | 22.0 - 34.5 |
| Fat Mass (kg) | 11 | 250 | 19.3 ± 2.1 | 19.0 | 15.1 - 22.9 | - | - | - | - | - |
| Fat-Free Mass (kg) | 12 | 518 | 37.8 ± 3.1 | 40.2 | 19.1– 40.2 | - | - | - | - | - |
| BMD (g/cm<sup>2</sup>) |  |  |  |  |  |  |  |  |  |  |
| Femoral Neck | 7 | 1146 | 0.84 ± 0.01 | 0.84 | 0.83 – 0.86 | 2 | 47 | 0.94± 0.01 | 0.94 | 0.93 – 0.95 |
| Lumbar Spine | 14 | 1344 | 1.09 ± 0.07 | 1.09 | 0.96 – 1.22 | 3 | 67 | 1.11 ± 0.08 | 1.11 | 1.01 - 1.21 |
| Total Hip | 10 | 1275 | 0.98 ± 0.04 | 0.95 | 0.95 – 1.07 | - | - | - | - | - |

Notes: #, Number of; SD, Standard Deviation; Mdn, Median; y, years; BMI, Body Mass Index; BMD, Bone Mineral Density; g/cm, grams per centimetre.

### GAHT Intervention Characteristics

A description of the GAHT interventions can be seen in **Table 2**. The GAHT interventions varied in length from 1 to 26 years (mean ± SD = 4 ± 5, median 2 years). Compliance with GAHT was not measured in any study. Seven groups accounted for physical activity, fourteen groups consumed alcohol, and twenty-five groups included smokers. One group contained patients with osteoporosis, and one group included patients using glucocorticoids. No groups presented with osteoarthritis or sarcopenia.

### BMD Assessment Characteristics

All studies assessing FN, TH, or LS BMD used DXA. Fourteen studies used a Hologic DXA system (Discovery (Auer et al., 2020; Dobrolińska et al., 2019; Van Caenegem & TLSjoen, 2015; Vlot et al., 2019; Wiepjes et al., 2020; Wiepjes et al., 2019; Wiepjes et al., 2017), horizon (B. Hamilton et al., 2024), or 49-159 (C. Pelusi et al., 2014), while two studies used the Norland DXA system (A. Turner et al., 2004; P. van Kesteren et al., 1998). One study did not report the DXA model (Iwamoto et al., 2024). Insufficient data were reported for the site-specific reliability of the instruments used to assess BMD at either the LS or FN. One study utilised a pQCT device (XCT-2000 (Van Caenegem & TLSjoen, 2015))

### Muscle Assessment Characteristics

Two studies measured muscle Cross-sectional Area (mCSA), where one utilised a pQCT device (XCT-2000 (Van Caenegem & TLSjoen, 2015). One study measured muscular power, with one utilising a JUM001 Jump Mat (Hamilton et al., 2021). Four studies measured muscular strength via handgrip (Blair Hamilton et al., 2024; Scharff et al., 2019; Y. Tominaga et al., 2025; Van Caenegem & TLSjoen, 2015), one study utilised Isokinetic dynamometry (T. R. Lundberg et al., 2024) and two utilised military fitness tests for strength (Chiccarelli et al., 2023; Roberts et al., 2021).

### Risk of Bias Assessment

Assessment using the ROBINS-I (J. A. Sterne et al., 2016) is shown in **Figure 2** and the **Supplementary Material file RoB (Brown et al., 2024a)** while the JBI Cross-Sectional checklist can be found in the **Supplementary file JBI_Checklist (Brown et al., 2024a)** . As can be seen, 45% of the studies were at a low risk of bias, 23% at a moderate risk of bias, 9% at a serious risk of bias and 24% at a critical risk of bias. Given the inability to blind participants in GAHT intervention trials, all longitudinal studies were judged at moderate risk of bias in the domain of deviations from intended interventions.” The overall risk of bias across all categories was categorised as moderate. All included cross-sectional studies met 6 out of the 8 criteria on the JBI. The JBI checklist showed that 75% of cross-sectional studies did not have strategies to deal with confounding factors (Brown et al., 2024a).

**Figure 2.**
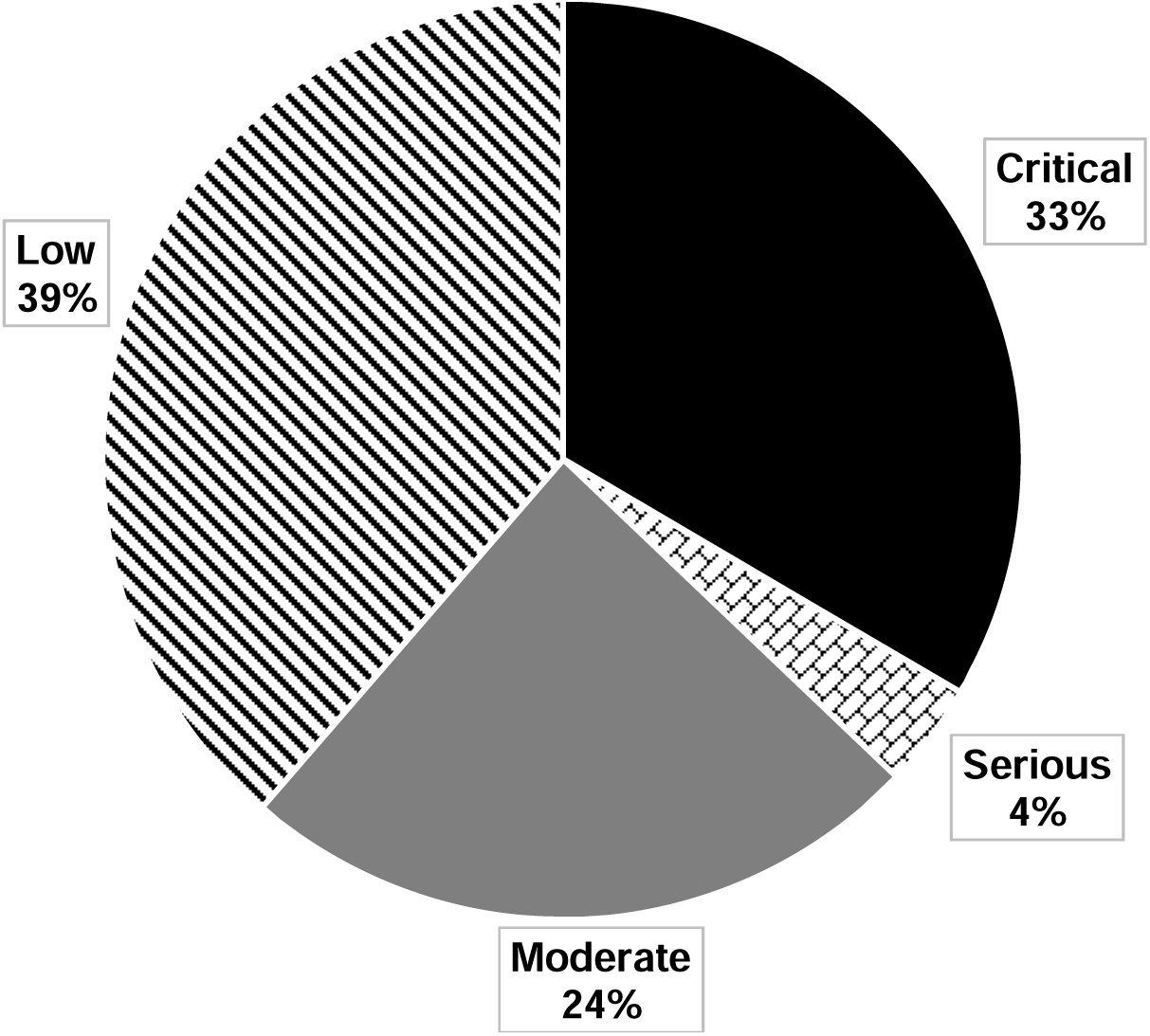
Risk Of Bias In Non-Randomized Studies - of Interventions (ROBINS-I)

### Primary Outcome Measures

#### FN BMD

Longitudinal Changes in FN BMD in transgender men with GAHT can be seen in **Table 4** and **Figure 3a**. Longitudinally, no change in FN BMD was found with GAHT (*g* = -0.10 [-0.41, 0.22], Z = -0.61, p = 0.54), with no asymmetry (LFK index 0.33) observed. In addition, statistically significant heterogeneity was observed (Q = 26.67, p = 0.00), and inconsistency was considered very high (*I^2^* = 78%). An evidence profile for changes in FN BMD is shown in **Supplementary Table 1**. Based on GRADE, the evidence was considered low quality, with future additional studies potentially influencing the overall direction of findings. Cross-sectional comparisons are seen in **Table 4** and **Figure 3b**. Transgender men compared with cisgender women showed an observed statistically significant difference in FN BMD (g = 1.00 [0.22, 1.77], Z = 2.52, p = 0.01) in favour of cisgender women, with no statistically significant heterogeneity observed (Q = 2.96, p = 0.90), though inconsistency was moderate (*I^2^* = 66).

**Figure 3.**
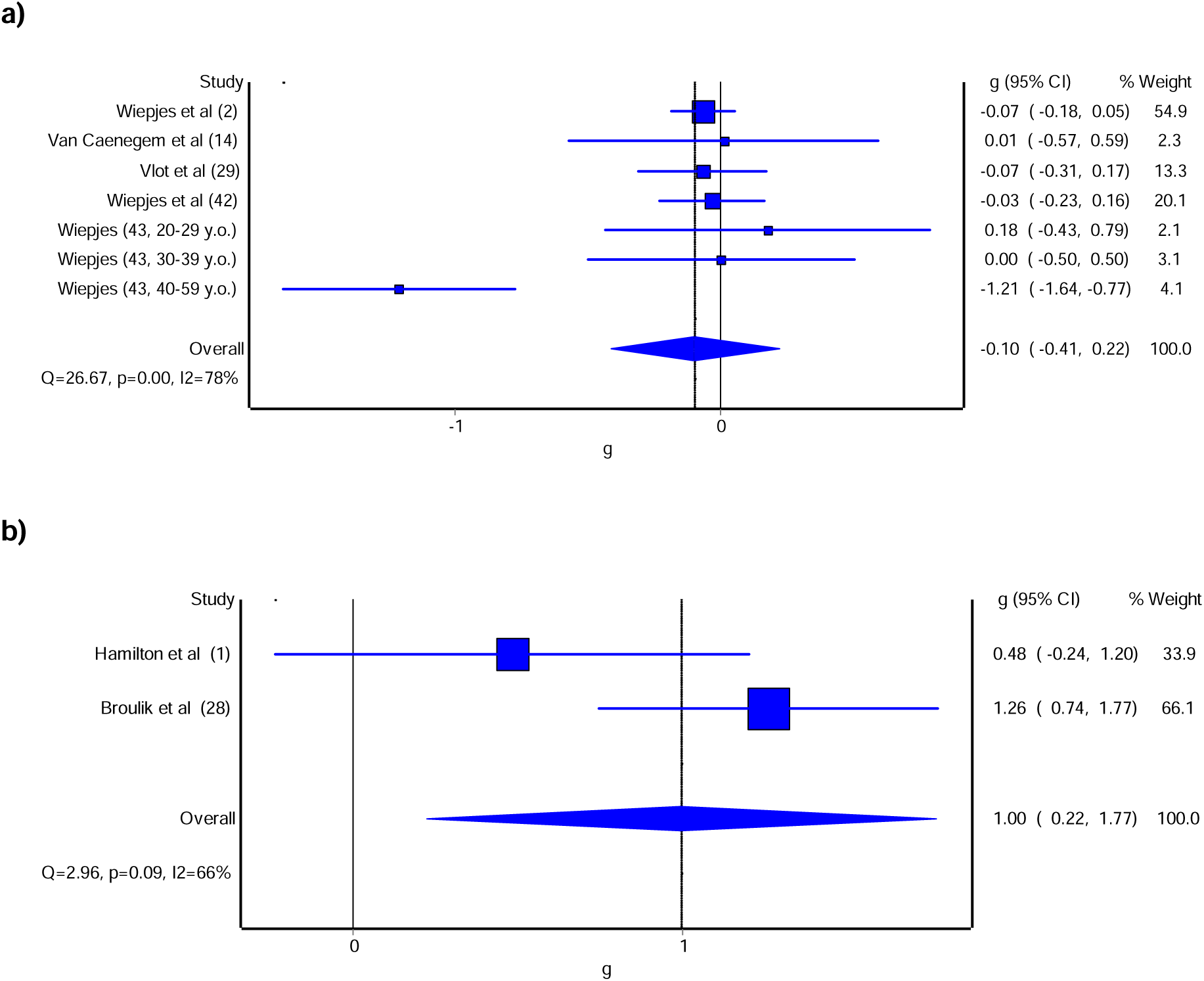
Forest plot for longitudinal. a) and cross-sectional comparisons b) in transgender men and cross-sectional comparisons b) of FN BMD. Forest plot for point estimates, standardised effect size changes (g) in FN BMD. The blue squares represent the standardised mean difference (g) while the left and right extremes of the squares represent the corresponding 95% confidence intervals. The middle of the blue diamond represents the overall standardised mean difference (g) while the left and right extremes of the diamond represent the corresponding 95% confidence intervals.

**Table 4.**
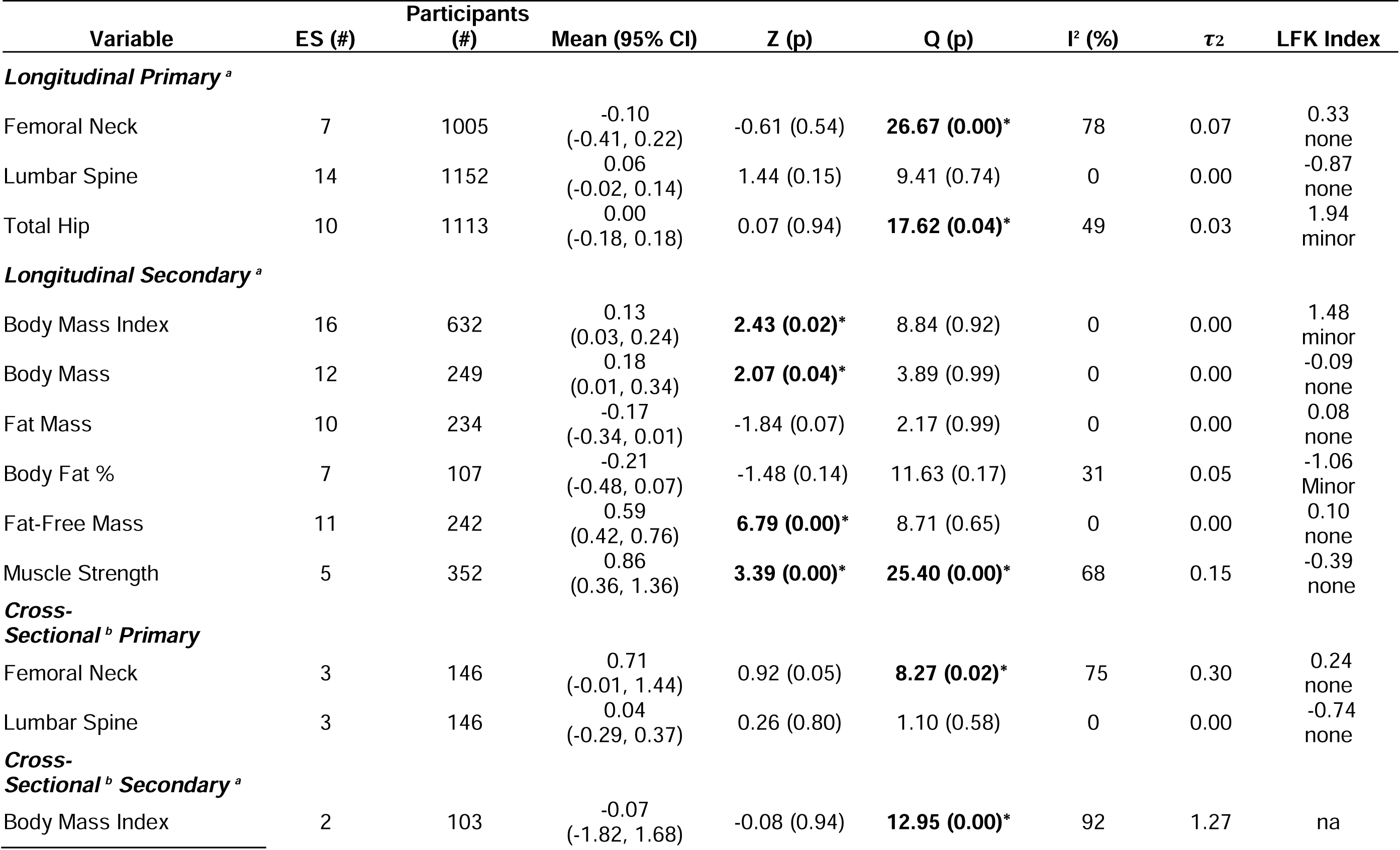
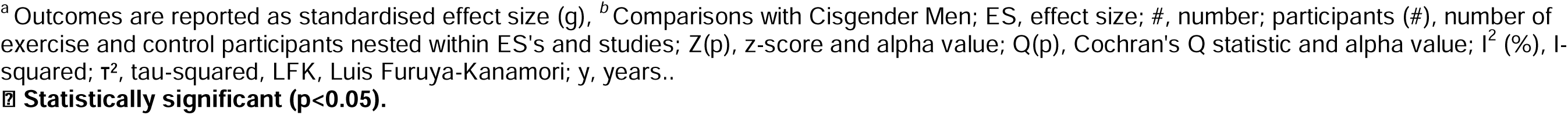
Results of primary and secondary outcomes.

| Variable | ES (#) | Participants<br>(#) | Mean (95% CI) | Z (p) | Q (p) | I<sup>2</sup> (%) | $\tau^2$ | LFK Index |
| --- | --- | --- | --- | --- | --- | --- | --- | --- |
| ***Longitudinal Primary<sup>a</sup>*** |  |  |  |  |  |  |  |  |
| Femoral Neck | 7 | 1005 | -0.10<br>(-0.41, 0.22) | -0.61 (0.54) | **26.67 (0.00)\*** | 78 | 0.07 | 0.33<br>none |
| Lumbar Spine | 14 | 1152 | 0.06<br>(-0.02, 0.14) | 1.44 (0.15) | 9.41 (0.74) | 0 | 0.00 | -0.87<br>none |
| Total Hip | 10 | 1113 | 0.00<br>(-0.18, 0.18) | 0.07 (0.94) | **17.62 (0.04)\*** | 49 | 0.03 | 1.94<br>minor |
| ***Longitudinal Secondary<sup>a</sup>*** |  |  |  |  |  |  |  |  |
| Body Mass Index | 16 | 632 | 0.13<br>(0.03, 0.24) | **2.43 (0.02)\*** | 8.84 (0.92) | 0 | 0.00 | 1.48<br>minor |
| Body Mass | 12 | 249 | 0.18<br>(0.01, 0.34) | **2.07 (0.04)\*** | 3.89 (0.99) | 0 | 0.00 | -0.09<br>none |
| Fat Mass | 10 | 234 | -0.17<br>(-0.34, 0.01) | -1.84 (0.07) | 2.17 (0.99) | 0 | 0.00 | 0.08<br>none |
| Body Fat % | 7 | 107 | -0.21<br>(-0.48, 0.07) | -1.48 (0.14) | 11.63 (0.17) | 31 | 0.05 | -1.06<br>Minor |
| Fat-Free Mass | 11 | 242 | 0.59<br>(0.42, 0.76) | **6.79 (0.00)\*** | 8.71 (0.65) | 0 | 0.00 | 0.10<br>none |
| Muscle Strength | 5 | 352 | 0.86<br>(0.36, 1.36) | **3.39 (0.00)\*** | **25.40 (0.00)\*** | 68 | 0.15 | -0.39<br>none |
| ***Cross-Sectional<sup>b</sup> Primary*** |  |  |  |  |  |  |  |  |
| Femoral Neck | 3 | 146 | 0.71<br>(-0.01, 1.44) | 0.92 (0.05) | **8.27 (0.02)\*** | 75 | 0.30 | 0.24<br>none |
| Lumbar Spine | 3 | 146 | 0.04<br>(-0.29, 0.37) | 0.26 (0.80) | 1.10 (0.58) | 0 | 0.00 | -0.74<br>none |
| ***Cross-Sectional<sup>b</sup> Secondary<sup>a</sup>*** |  |  |  |  |  |  |  |  |
| Body Mass Index | 2 | 103 | -0.07<br>(-1.82, 1.68) | -0.08 (0.94) | **12.95 (0.00)\*** | 92 | 1.27 | na |

<sup>a</sup> Outcomes are reported as standardised effect size (g), <sup>b</sup> Comparisons with Cisgender Men; ES, effect size; #, number; participants (#), number of exercise and control participants nested within ES's and studies; Z(p), z-score and alpha value; Q(p), Cochran's Q statistic and alpha value; I<sup>2</sup> (%), I- squared; τ<sup>2</sup>, tau-squared, LFK, Luis Furuya-Kanamori; y, years.. **■ Statistically significant (p<0.05).**

#### LS BMD

Longitudinal changes in LS BMD in transgender men with GAHT can be seen in **Table 4** and **Figure 4a**. There was no observed statistically significant change with GAHT on LS BMD (g = 0.06 [-0.02, 0.14], Z = 1.44, p = 0.15), with no asymmetry (LFK index -0.87) observed. In addition, heterogeneity was not statistically significant (Q = 9.41, p = 0.74), and inconsistency was low (*I^2^* = 0%). Findings were similar when results were collapsed so that only one effect size represented each study. With the removal of each effect size from the model, the results remained non-significant. An evidence profile for changes in LS BMD is shown in **Supplementary Table 1**. Based on GRADE, the evidence was considered low quality, with future additional studies likely to influence the overall direction of findings. Transgender men compared with cisgender women cross-sectionally showed no observed statistically significant difference in LS BMD (g = 0.15 [-0.24, 0.55], Z = 0.77, p = 0.44).

**Figure 4.**
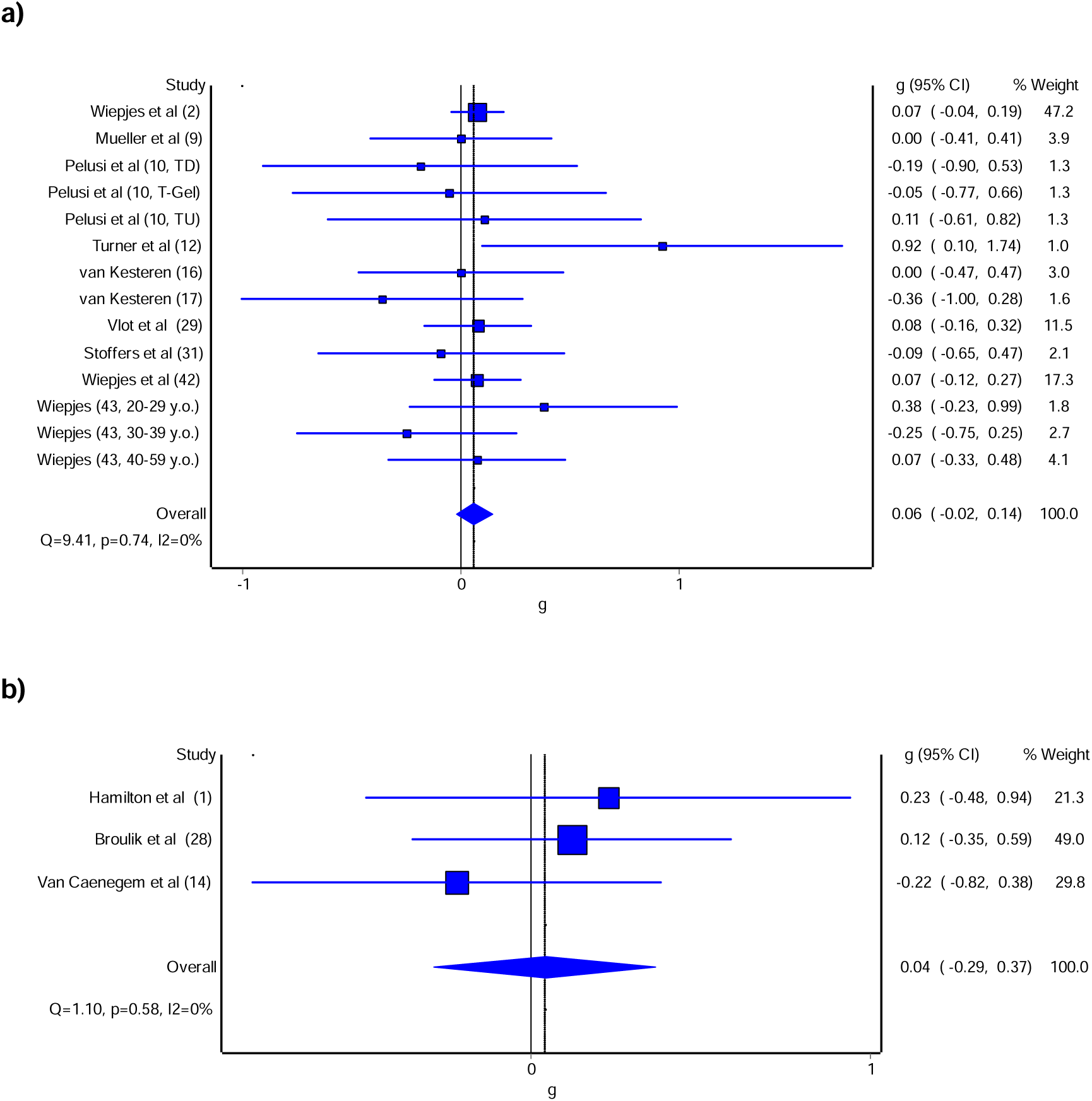
Forest plot for longitudinal. a) and cross-sectional comparisons b) changes with GAHT in transgender men of LS BMD. The blue squares represent the standardised mean difference (g) while the left and right extremes of the squares represent the corresponding 95% confidence intervals. The middle of the blue diamond represents the overall standardised mean difference (g) while the left and right extremes of the diamond represent the corresponding 95% confidence intervals.

#### TH BMD

Longitudinal changes in TH BMD in transgender men with GAHT can be seen in **Table 4** and **Figure 5**. There was no observed statistically significant change with GAHT on TH BMD (g = 0.00 [-0.18, 0.18], Z =0.07, p = 0.94), with minor asymmetry (LFK index = 1.94 observed. Heterogeneity was statistically significant (Q = 17.62, p = 0.04), and inconsistency was moderate (*I^2^*= 49%). No removal of any studies affected the analysis (**Supplementary Table 3**). An evidence profile for changes in TH BMD is shown in online **Supplementary Table 1**. Based on GRADE, the evidence was considered low quality, with future additional studies potentially influencing the overall direction of findings.

**Figure 5.**
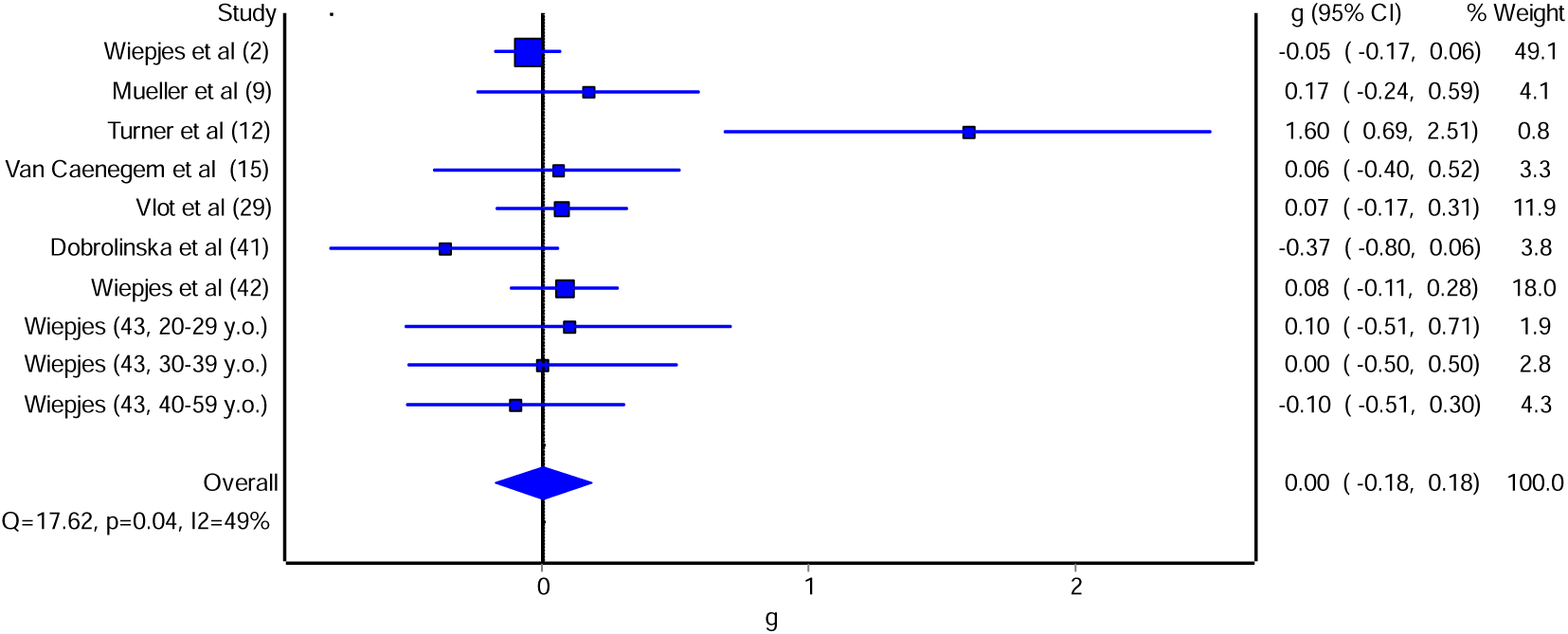
Forest plot for longitudinal changes with GAHT in transgender men of TH BMD. Forest plot for point estimates standardised effect size changes (g) in TH BMD. The blue squares represent the standardised mean difference (g) while the left and right extremes of the squares represent the corresponding 95% confidence intervals. The middle of the blue diamond represents the overall standardised mean difference (g) while the left and right extremes of the diamond represent the corresponding 95% confidence intervals.

### Changes in Secondary Outcomes

#### Body Mass Index (BMI)

Longitudinal changes in BMI in transgender men with GAHT can be seen in **Table 4** and **Figure 6a**. There was an observed statistically significant effect of GAHT on BMI (g = 0.13 [0.03, 0.24], Z = 2.43, p = 0.02), with minor asymmetry (LFK index = 1.48) observed. In addition, no statistically significant heterogeneity was observed (Q = 8.84, p = 0.92), and overall inconsistency was considered to be low (*I^2^* = 0%). Influence analysis showed that the removal of other studies from the analysis did not change the outcome. Based on GRADE, the evidence was considered moderate, with future additional studies potentially influencing the overall direction of findings. Transgender men compared with cisgender women cross-sectionally showed no observed statistically significant difference in BMI (g = -0.07 [-1.82, 1.68], Z = -0.08, p = 0.94, **Figure 6b**). An evidence profile for changes in BMI is shown in **Supplementary Table 1.**

**Figure 6.**
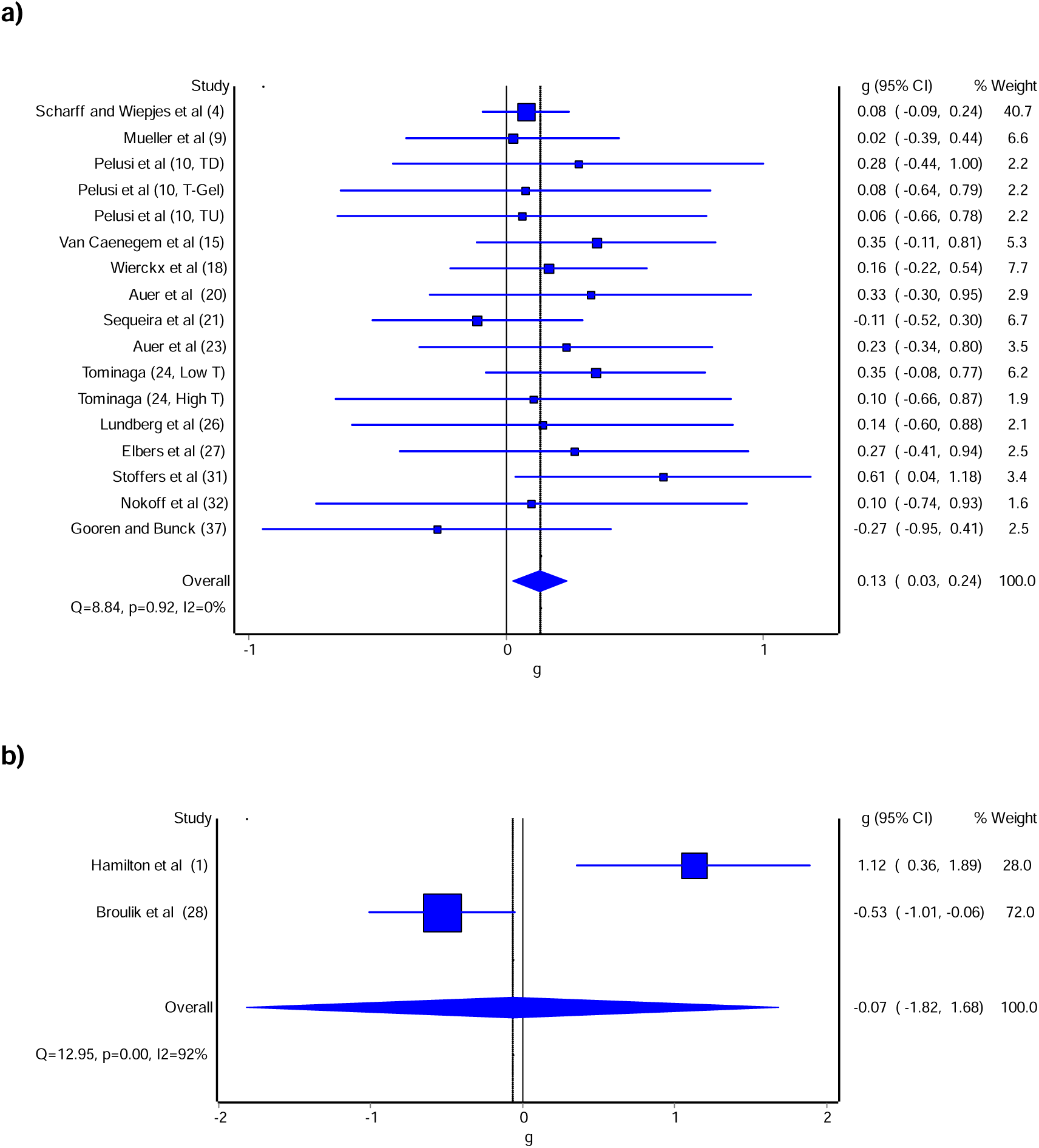
Forest plot for longitudinal changes. a) and cross-sectional comparisons b) with GAHT in transgender of BMI. Forest plot for point estimate standardised effect size changes (g) BMI. The blue squares represent the standardised mean difference (g) while the left and right extremes of the squares represent the corresponding 95% confidence intervals. The middle of the blue diamond represents the overall standardised mean difference (g) while the left and right extremes of the diamond represent the corresponding 95% confidence intervals.

#### Body Mass (BM)

Longitudinal changes of BM in transgender men with GAHT can be seen in **Table 4** and **Figure 7a**. There was a statistically significant effect of GAHT on BM (g = 0.18 [0.01, 0.34], Z = 2.07, p = 0.04), with no asymmetry (LFK index = -0.09) observed. In addition, no statistically significant heterogeneity was observed (Q = 2.17, p = 0.99), and overall inconsistency was low (*I^2^* = 0%). Influence analysis indicated that exclusion of the Van Caenegem *et al*., the result became statistically significant (**Supplementary Table 2**). Based on GRADE, the evidence was considered moderate, with future additional studies potentially influencing the overall direction of findings (**Supplementary Table 1**).

**Figure 7.**
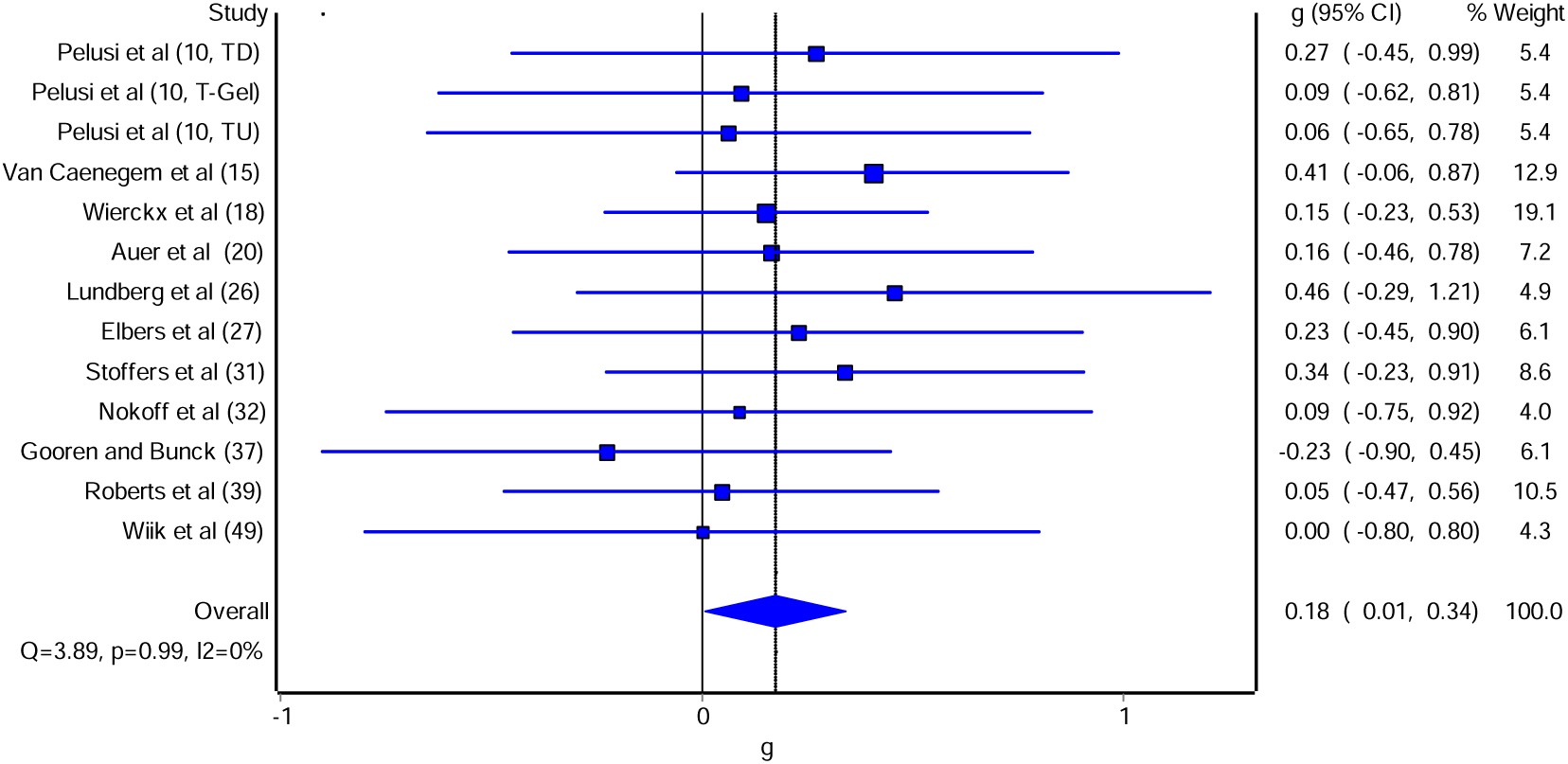
Forest plot for longitudinal changes with GAHT in transgender men of Body Mass. The blue squares represent the standardised mean difference (g) while the left and right extremes of the squares represent the corresponding 95% confidence intervals. The middle of the blue diamond represents the overall standardised mean difference (g) while the left and right extremes of the diamond represent the corresponding 95% confidence intervals.

#### Fat Mass (FM)

Longitudinal Results of FM in transgender men with GAHT can be seen in **Table 4** and **Figure 8a**. A statistically significant effect of GAHT on FM (g = -0.17 [-0.34, 0.01], Z = -1.84, p = 0.07), with minor asymmetry (LFK index -0.43), **Supplementary figure 6a**) was observed. In addition, no statistically significant heterogeneity was observed (Q = 2.17, p = 0.99), and overall inconsistency was very low (*I^2^* = 0%). Influence analysis showed that the removal of any study did not change the outcome. Based on GRADE, the evidence was considered high quality and further research is unlikely to significantly change the confidence in the estimate (**Supplementary Table 1**).

**Figure 8.**
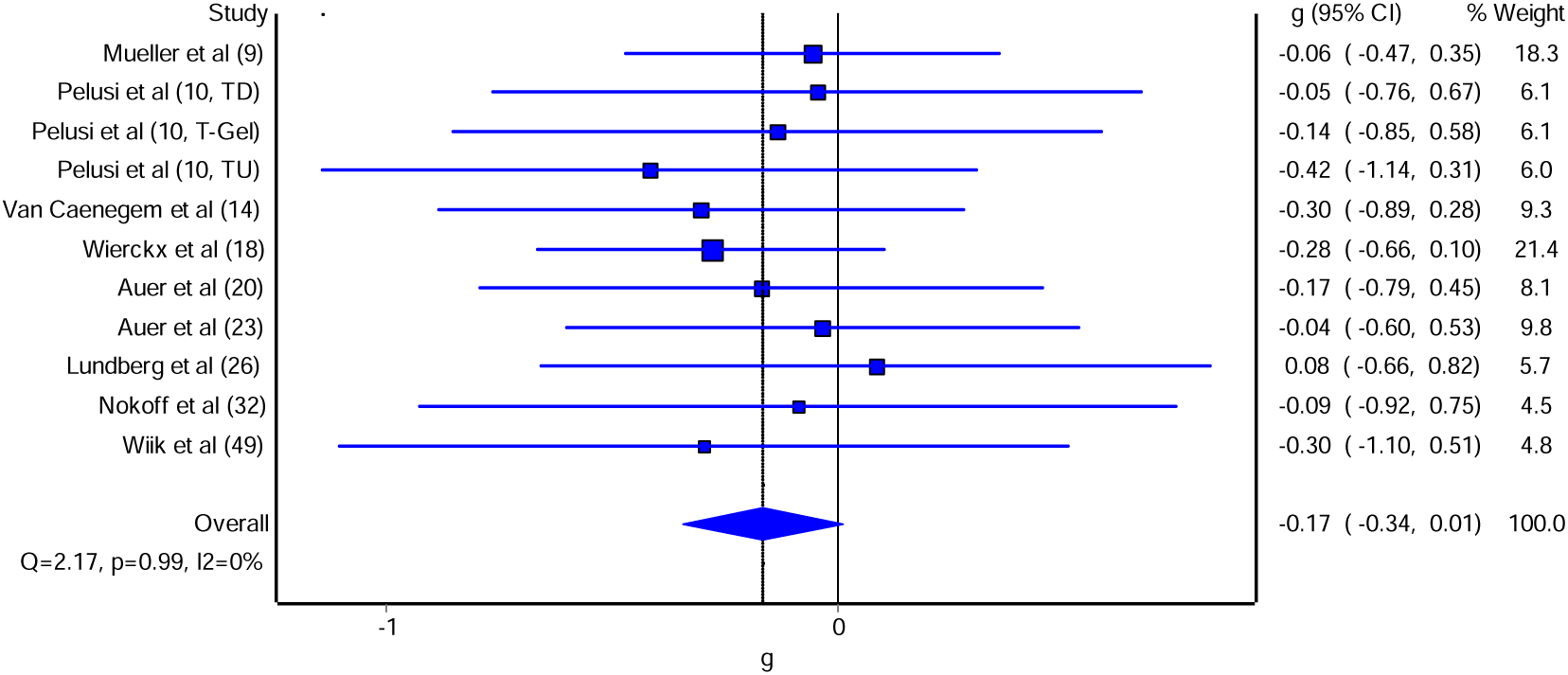
Forest plot for longitudinal changes with GAHT in transgender men of Fat Mass. The blue squares represent the standardised mean difference (g) while the left and right extremes of the squares represent the corresponding 95% confidence intervals. The middle of the blue diamond represents the overall standardised mean difference (g) while the left and right extremes of the diamond represent the corresponding 95% confidence intervals.

#### Body Fat % (BF%)

Longitudinal Results of BF% in transgender men with GAHT can be seen in **Table 4** and **Figure 9a**. There was no significant effect of GAHT on BF% (g = -0.21 [-0.48, 0.07], Z = - 1.48, p = 0.14), with minor asymmetry (LFK index -1.06) observed. In addition, no significant heterogeneity was observed (Q = 11.63, p = 0.17), and overall inconsistency was considered to be low (*I^2^* = 31%). Influence analysis indicated that exclusion of the Tominaga *et al*., low testosterone groups the result became statistically significant (**Supplementary Table 3**). Based on GRADE, the evidence was considered moderate, with future additional studies potentially influencing the overall direction of findings (**Supplementary Table 1**).

**Figure 9.**
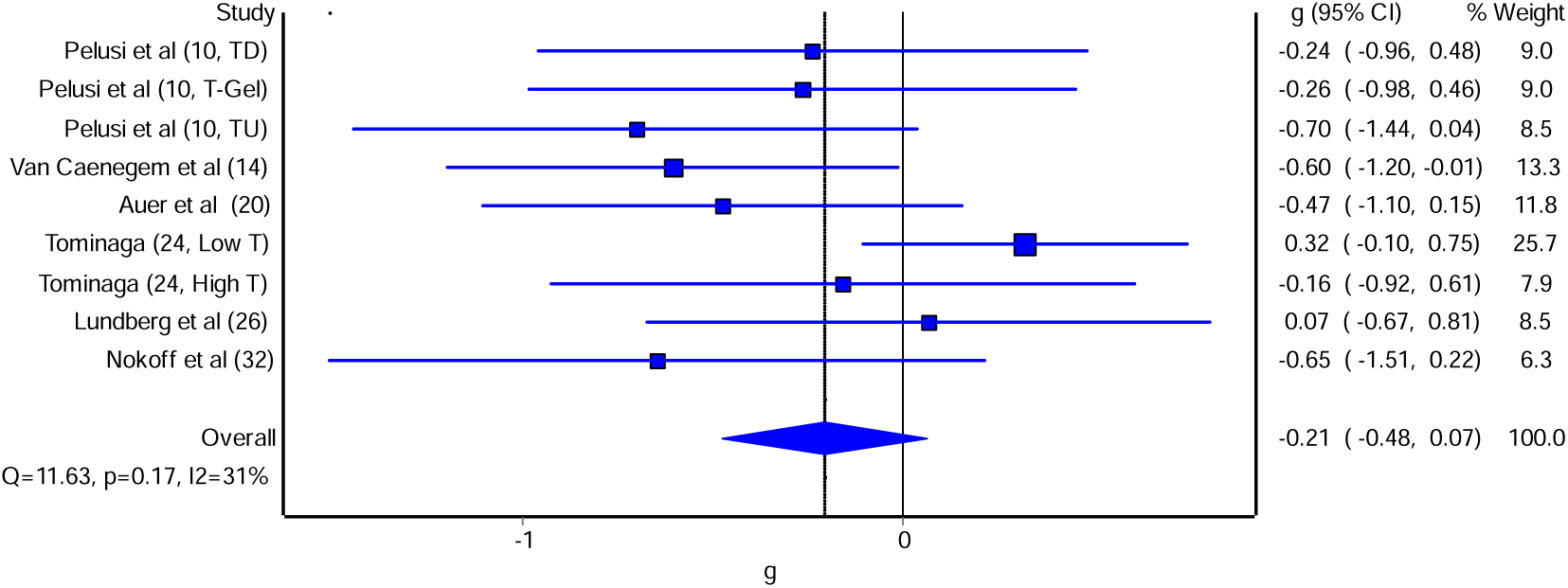
Forest plot for longitudinal changes with GAHT in transgender men of Body Fat %. The blue squares represent the standardised mean difference (g) while the left and right extremes of the squares represent the corresponding 95% confidence intervals. The middle of the blue diamond represents the overall standardised mean difference (g) while the left and right extremes of the diamond represent the corresponding 95% confidence intervals.

#### Fat-Free Mass (FFM)

Longitudinal Results of FFM in transgender men with GAHT can be seen in **Table 4** and **Figure 10a**. There was an observed statistically significant effect of GAHT on FFM (g = 0.59 [0.42, 0.76, Z = 6.79, p = 0.00), with no asymmetry (LFK index = 0.10) observed. In addition, no heterogeneity was observed (Q = 8.71, p = 0.65), and overall inconsistency was very low (*I^2^* = 0%). Influence analysis showed that the removal of any study did not change the outcome. Based on GRADE, the evidence was considered high quality, with future additional studies not influencing the overall direction of findings (**Supplementary Table 1**).

**Figure 10.**
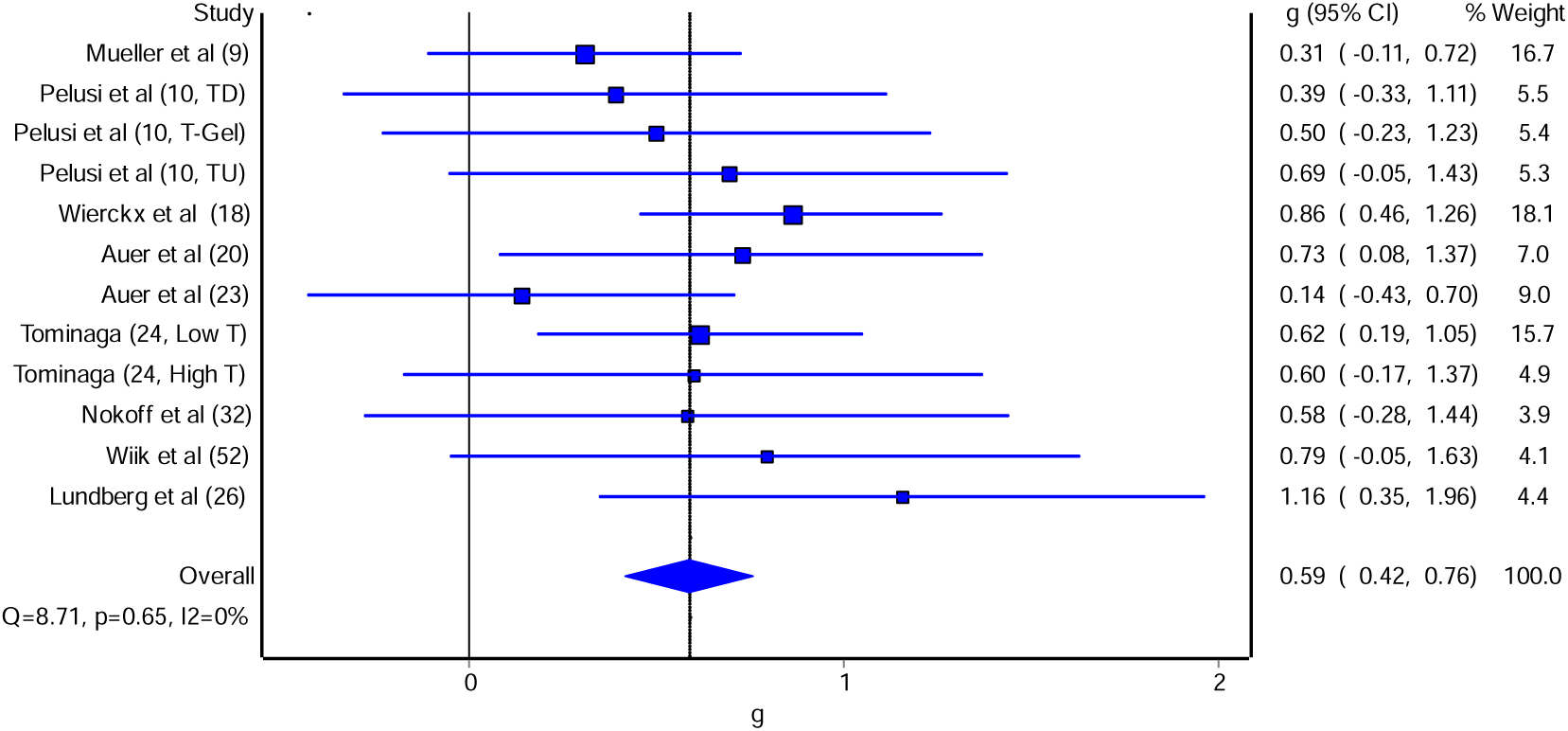
Forest plot for longitudinal changes with GAHT in transgender men of Fat Free Mass. The blue squares represent the standardised mean difference (g) while the left and right extremes of the squares represent the corresponding 95% confidence intervals. The middle of the blue diamond represents the overall standardised mean difference (g) while the left and right extremes of the diamond represent the corresponding 95% confidence intervals.

#### Muscle Strength

Longitudinal Results of Muscle Strength in transgender men with GAHT can be seen in **Table 4** and **Figure 11**. There was an observed statistically significant effect of GAHT on MCSA (g = 0.86 [0.36, 1.36] Z = 3.39 p = 0.00), with no asymmetry (LFK index= -0.39) observed. In addition, statistically significant heterogeneity was observed (Q = 25.4, p = 0.00), and overall inconsistency was moderate (I2 = 68%). Based on GRADE, the evidence was considered low quality, with future additional studies likely to influence the overall direction of findings (**Supplementary Table 1**).

**Figure 11.**
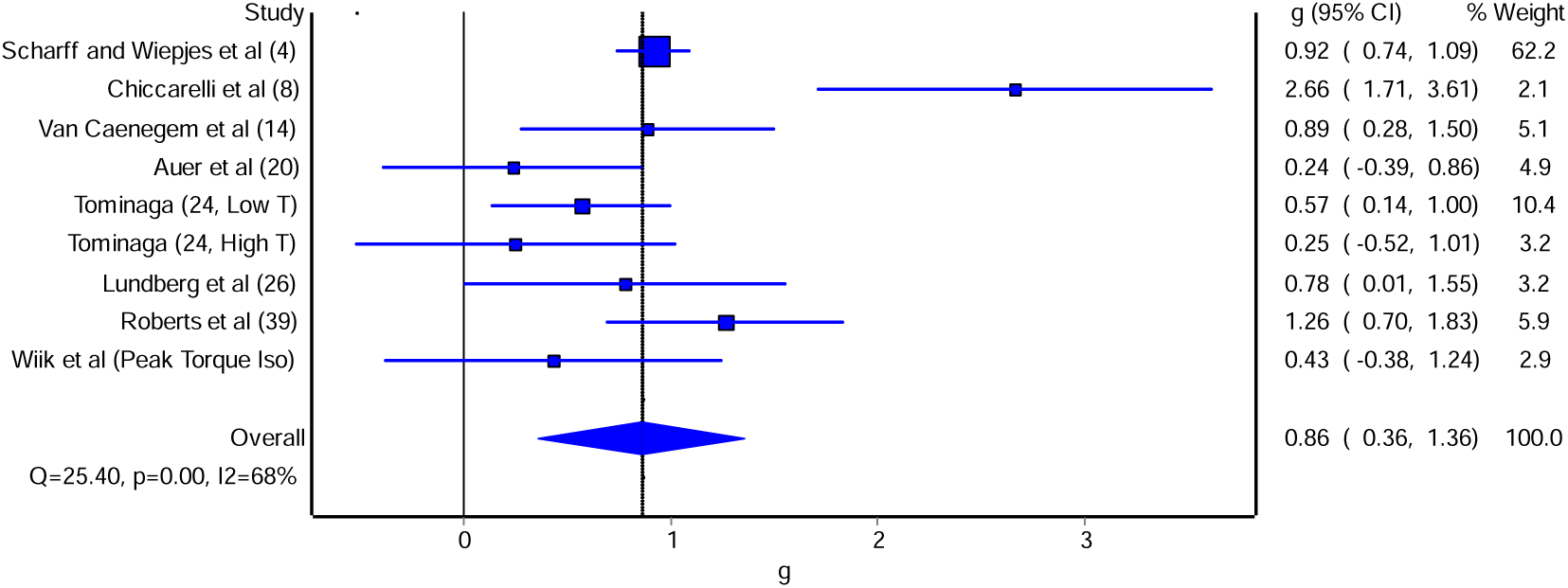
Forest plot for longitudinal changes with GAHT in transgender women of Muscle Strength. The blue squares represent the standardised mean difference (g) while the left and right extremes of the squares represent the corresponding 95% confidence intervals. The middle of the blue diamond represents the overall standardised mean difference (g) while the left and right extremes of the diamond represent the corresponding 95% confidence intervals.

## Discussion

The primary aim of the current systematic review and meta-analysis was to update the work by Singh-Ospina et al. (Singh-Ospina et al., 2017) examining the effects of GAHT on the Bone Health of Transgender men. The BMD findings suggest that an average of 5 years of masculine GAHT is not associated with changes in LS, FN, or TH BMD, consistent with findings from (Singh-Ospina et al., 2017). Peak bone mass in individuals assigned female at birth is typically reached by approximately ages 20-30 and plateaus until age 50, after which bone mass begins to decrease (Hereford et al., 2024; Lu et al., 2016). The transgender men in this study had a mean age of 28 ± 6 years; therefore, the influence of age-related bone gains or losses is expected to be minimal, supporting the validity of the findings presented. However, one of the most striking findings of this meta-analysis is the consistently low quality of evidence underpinning bone-related outcomes in transgender men undergoing GAHT. Across all assessed bone metrics, the evidence base was characterised by marked publication bias, moderate to high statistical inconsistency, and notable risks of bias, alongside substantial indirectness due to the heterogeneity of GAHT regimens and methodologies used across studies. Although Singh-Ospina et al. (Singh-Ospina et al., 2017) and Fighera et al (Tayane Muniz Fighera et al., 2019) both report a neutral effect of GAHT on LS, FN, or TH BMD at 12 or 24 months of follow-up; such conclusions cannot be considered robust given the underlying methodological weaknesses and severe imprecision shown in this meta-analysis (**Table 4**). Our sister meta-analysis in transgender women demonstrated that the quality of evidence for two of the three clinical sites, the FN and TH, was at least of moderate quality (Andrew Brown et al., 2025), indicating a notable disparity in the strength of evidence between transgender women and transgender men. This gap in the evidence base is likely to have meaningful implications for clinical decision-making and long-term healthcare provision for transgender men and therefore must be addressed urgently. Our findings highlight the need for well-designed, adequately powered clinical trials for bone health that directly confront current sources of bias, particularly regimen variability, short follow-up durations, and inconsistent reporting, to enable a clearer understanding of the true effects of differing GAHT treatment protocols on BMD trajectories.

When focusing specifically on the cross-sectional comparisons of bone mineral density, both our meta-analysis and that of Fighera et al (Tayane Muniz Fighera et al., 2019) showed only trivial differences between transgender men and cisgender women. At the femoral neck, Fighera et al.(Tayane Muniz Fighera et al., 2019) reported a very small mean difference (MD = 0.05) in favour of transgender men with high heterogeneity (I² = 85%), while our standardised analysis produced an equally small effect in the opposite direction, favouring cisgender women (*g* = –0.10) with similarly substantial heterogeneity (I² = 75%). Singh-Ospina et al. (Singh-Ospina et al., 2017) did not perform a cross-sectional analysis between transgender men and cisgender women for bone health. These minimal and directionally inconsistent point estimates likely reflect the small number of eligible cross-sectional studies and substantial variability in study populations and measurement methods. At the lumbar spine, both reviews again identified negligible differences (MD = – 0.02; *g* = 0.06) with low heterogeneity (I² = 19% and 0%, respectively). Collectively, the cross-sectional evidence base, only three studies in our analysis (*n* = 146 transgender men = 79), highlights how poorly this comparison is currently understood and underscores the need for more rigorously designed studies and work in this area.

Our secondary aim was to update the work of Singh-Ospina et al. (Singh-Ospina et al., 2017) and extend the analysis to overall musculoskeletal health, bringing insights into GAHT’s longitudinal effects in transgender men. The overall findings suggest that longitudinal GAHT is associated with statistically significant gains in BMI, BM, FFM and muscle strength (**Table 4**). GAHT was not associated with a significant overall decline in BF%; however, this finding was sensitive to influence analysis. The original model showed moderate heterogeneity and a non-significant pooled effect. After removing the study by Tominaga et al (Y. Tominaga et al., 2025), the effect became a statistically significant decline in BF%, accompanied by the elimination of heterogeneity (**Supplementary Table 3**), indicating that the original null finding was strongly influenced by the low dose of masculine GAHT in Tominaga et al (Y. Tominaga et al., 2025). Body mass changes were also sensitive to influence analysis after removing Van Caenegem et al. (2015) (Van Caenegem et al., 2015), resulting in a non-significant change in BM (**Supplementary Table 3)** once this study was removed. These sensitivities further illustrate that the field remains in an early stage of empirical development, and current conclusions must be regarded as provisional until confirmed by higher-quality, replicated longitudinal research in musculoskeletal health.

Longitudinal evidence indicates that GAHT is associated with increases in muscle strength (**Table 4**), although these findings are accompanied by moderate inconsistency and heterogeneity. Much of this variability appears to stem from the use of three distinct assessment modalities, handgrip dynamometry, isokinetic dynamometry, and functional fitness testing, with an overreliance on handgrip strength. Handgrip performance should be interpreted cautiously in transgender men, as it is strongly influenced by anthropometric factors such as finger length, finger span, and hand perimeter, none of which are modified by GAHT; consequently, this measure may underestimate the true extent of strength gains (Cheung et al., 2024) (Cheung et al., 2024). Notably, our meta-analysis found that testosterone-based GAHT was associated with concurrent increases in fat-free mass and muscle strength. This pattern contrasts with (Zhang et al., 2025) (Zhang et al., 2025), who reported that testosterone levels are positively associated with muscle mass but not muscle strength in cisgender young to middle-aged males, suggesting that transgender men assigned female-at-birth may exhibit greater sensitivity to the anabolic and neuromuscular effects of exogenous testosterone. While longitudinal transmasculine GAHT elicits biologically coherent increases in body composition parameters (**Table 4**), in contrast to the reductions in fat-free mass observed in transgender women following testosterone suppression and oestrogen supplementation in our sister meta-analysis (Andrew Brown et al., 2025), the direct anabolic effects of exogenous testosterone (Bhasin et al., 2025) in transgender men are expected to produce substantial gains in muscle strength that are unlikely to be fully mitigated by low physical activity. The heterogeneity in strength outcomes, therefore, likely reflects methodological inconsistency in the method rather than physiological ambiguity. To better delineate muscle strength trajectories in transgender men receiving GAHT, further longitudinal research using standardised strength assessments and well-matched comparator groups is critically needed.

While transgender men exhibit lower BMI than cisgender women in cross-sectional analyses, this is plagued by significant heterogeneity and high inconsistency. This discrepancy underscores the need for caution when interpreting cross-sectional data, especially since none of the cross-sectional data from our analysis was of matched cohorts. Further research with well-matched comparators is critical to clarify muscle BMI trajectories in transgender men undergoing GAHT, together with the significant results of the influence analysis (Supplementary Table 4).

### Limitations

Another important limitation in the current body of literature, and consequently in our review, is the predominant reliance on DXA as the imaging modality for assessing bone health. While DXA provides clinically useful measures of areal bone mineral density (aBMD), it does not capture bone microarchitecture or strength parameters, which are critical determinants of fracture risk. High-resolution peripheral quantitative computed tomography (HR-pQCT), a more advanced imaging technique, offers superior resolution of trabecular and cortical microstructure and may serve as a better surrogate for fracture risk, particularly in populations with altered bone physiology such as transgender men.

Emerging evidence from HR-pQCT studies provides important insight into skeletal microarchitecture in transgender men. Bretherton et al. (2022) (Bretherton et al., 2022) reported that, relative to cisgender women, transgender men demonstrated significantly higher total volumetric bone mineral density (vBMD; 0.63 SD, p = 0.01). Although cortical vBMD and cortical porosity did not differ between groups, cortical thickness was markedly greater in transgender men (1.11 SD, p < 0.01), and trabecular thickness was modestly but significantly higher (0.38 SD, p = 0.05), with no other trabecular parameters showing notable differences. These findings indicate that bone microarchitecture in transgender men is not compromised and may be preserved through the aromatisation of exogenous testosterone to oestradiol, which could play a protective role against bone loss. This underscores the need for more nuanced research into testosterone dosing, the extent of aromatisation to oestradiol, and the skeletal implications of both hormones. Future primary studies should prioritise the integration of HR-pQCT outcomes, in addition to DXA, where available and investigate the relationships among circulating hormone levels, bone microarchitecture, and fracture risk in greater depth. Addressing these gaps will be critical for optimising clinical management and improving long-term bone health outcomes in transgender men.

A key limitation of this meta-analysis is the lack of matching of groups for age/height/BMI or uniformity across studies regarding the type, duration, and dosing of GAHT, as well as the variable inclusion of gonadectomy status. These factors are known to significantly influence musculoskeletal outcomes, particularly BMD and long-term bone health. Additionally, nutrition, physical activity, differences in age at initiation of GAHT and duration of hormone exposure introduce further heterogeneity, potentially impacting the comparability of findings across studies and limiting the precision of pooled estimates.

### Implications

This meta-analysis provides several clinically relevant insights for the management of transgender men undergoing GAHT. First, although BMD generally appears stable over approximately five years of therapy, the consistently low certainty of evidence and substantial methodological variability across studies indicate that clinicians should continue to conduct routine bone health monitoring, particularly at the femoral neck, and maintain vigilance in individuals with established risk factors for low bone mass. Second, the robust and consistent increases in fat-free mass suggest that masculinising GAHT confers meaningful anabolic benefits; clinicians should therefore reassure service users that increases in weight may reflect healthy gains in lean tissue, while simultaneously supporting engagement in resistance-based physical activity to optimise musculoskeletal adaptation. Third, the reliance on handgrip strength in existing studies highlights the need for gender clinics and researchers to come together to adopt more comprehensive strength assessments, particularly lower-limb functional testing, when evaluating musculoskeletal change during GAHT. Finally, emerging HR-pQCT evidence suggests preserved or enhanced bone microarchitecture, which underscores the importance of maintaining adequate testosterone dosing and may inform future refinement of hormonal monitoring. Together, these findings highlight the need for a proactive, individualised approach to musculoskeletal surveillance in transgender men and underscore the value of integrating standardised assessment protocols into clinical services until higher-quality longitudinal research becomes available.

## Conclusion

This systematic review demonstrates that masculinising GAHT produces clear anabolic effects on body composition and muscle strength, while available evidence suggests no significant longitudinal change in BMD. However, the consistently low certainty of bone-related evidence, high heterogeneity, and limited cross-sectional data highlight how poorly musculoskeletal health in transgender men is currently understood. The field now urgently requires rigorous, adequately powered longitudinal studies using standardised assessments and incorporating advanced imaging modalities. Until such data emerge, clinicians should maintain routine bone monitoring, support physical activity and strength development, and adopt an individualised approach to musculoskeletal care in transgender men

## Supporting information

JBI Checklist

Risk Of Bias

## Data Availability

All data produced are available online at Project DOI
10.17605/OSF.IO/42WY3

https://doi.org/10.17605/OSF.IO/42WY3

## Contributions

Conceptualisation, BRH; methodology, BRH; writing original draft preparation, BRH, KH, AB, SMM; Data Abstraction. AB, SMM, BRH; Risk of Bias, AB and SMM; GRADE, KH; JBI; KH, Clinical Applications, JS, LC, JB and BH; Data analysis, KH and BRH; writing--review and editing, ALL.

## Acknowledgements

The authors wish to express their gratitude to Dr Ferus Guppy for their guidance in developing the methodology and to Dr George Kelley for their guidance in the development of the codebook used for data abstraction. The authors would also like to thank Dr Tommy Lundberg for supplying the requested data from Wiik et al (2019).

## Abbreviations

The following abbreviations are used in this manuscript:

BMD: Bone Mineral Density
LM: Lean Mass
FFM: Fat-Free Mass
FM: Fat Mass
mCSA: Muscle Cross-Sectional Area
BMI: Body Mass Index
DPA: Dual-energy Photon Absorptiometry
DXA: Dual-energy X-ray Absorptiometry
ES: effect size
FN: Femoral Neck
TH: Total Hip
g: Hedges’ standardised mean difference effect size
GRADE: Grading of Recommendations Assessment, Development and Evaluation
IVhet: Inverse heterogeneity
LFK: Luis Furuya-Kanamori
LS: Lumbar Spine
NNS: Number-needed-to-screen
PRISMA: Preferred Reporting Items for Systematic reviews and Meta-Analyses
pQCT: peripheral Quantified Computer Tomography
SD: Standard Deviation

## References

Aguirre, J. I., Plotkin, L. I., Gortazar, A. R., Millan, M. M., O’Brien, C. A., Manolagas, S. C., & Bellido, T. (2007). A novel ligand-independent function of the estrogen receptor is essential for osteocyte and osteoblast mechanotransduction. J Biol Chem, 282(35), 25501–25508. 10.1074/jbc.M702231200

Ahn, S., & Becker, B. J. (2011). Incorporating Quality Scores in Meta-Analysis. Journal of Educational and Behavioral Statistics, 36(5), 555–585. 10.3102/1076998610393968

Aitken, M., Steensma, T. D., Blanchard, R., Vanderlaan, D. P., Wood, H., Fuentes, A., Spegg, C., Wasserman, L., Ames, M., Fitzsimmons, C. L., Leef, J. H., Lishak, V., Reim, E., Takagi, A., Vinik, J., Wreford, J., Cohen-Kettenis, P. T., De Vries, A. L. C., Kreukels, B. P. C., & Zucker, K. J. (2015). Evidence for an Altered Sex Ratio in Clinic-Referred Adolescents with Gender Dysphoria. The journal of sexual medicine, 12(3), 756–763. 10.1111/jsm.12817

Arcelus, J., Bouman, W. P., Van Den Noortgate, W., Claes, L., Witcomb, G., & Fernandez-Aranda, F. (2015). Systematic Review and Meta-Analysis of Prevalence Studies in Transsexualism. European Psychiatry, 30(6), 807–815. 10.1016/j.eurpsy.2015.04.005

Auer, M. K., Cecil, A., Roepke, Y., Bultynck, C., Pas, C., Fuss, J., Prehn, C., Wang-Sattler, R., Adamski, J., Stalla, G. K., & T’Sjoen, G. (2016). 12-months metabolic changes among gender dysphoric individuals under cross-sex hormone treatment: a targeted metabolomics study. Scientific reports, 6(1), 37005. 10.1038/srep37005

Auer, M. K., Ebert, T., Pietzner, M., Defreyne, J., Fuss, J., Stalla, G. K., & T’Sjoen, G. (2018). Effects of Sex Hormone Treatment on the Metabolic Syndrome in Transgender Individuals: Focus on Metabolic Cytokines. The Journal of Clinical Endocrinology & Metabolism, 103(2), 790–802. 10.1210/jc.2017-01559

Auer, M. K., Paizoni, L., Hofbauer, L. C., Rauner, M., Chen, Y., Schmidt, H., Huebner, A., Bidlingmaier, M., & Reisch, N. (2020). Effects of androgen excess and glucocorticoid exposure on bone health in adult patients with 21-hydroxylase deficiency. The Journal of steroid biochemistry and molecular biology, 204, 105734. 10.1016/j.jsbmb.2020.105734

Bhasin, S., Wang, C., Chandra, M. S., Gagliano-Jucá, T., & Jasuja, R. (2025). Mechanisms of Testosterone’s Anabolic Effects on Muscle and Function: Controversies and New Insights. Endocrine reviews. 10.1210/endrev/bnaf041

Bobba, R., Kalra, P., & Dharmalingam, M. (2023). Study of Effects of Gender-Affirming Hormone Therapy on Bone Mineral Density in Individuals with Gender Dysphoria. Indian J Endocrinol Metab, 27(6), 486–491. 10.4103/ijem.ijem_265_23

Bretherton, I., Ghasem-Zadeh, A., Leemaqz, S. Y., Seeman, E., Wang, X., McFarlane, T., Spanos, C., Grossmann, M., Zajac, J. D., & Cheung, A. S. (2022). Bone Microarchitecture in Transgender Adults: A Cross-Sectional Study [Article]. Journal of Bone & Mineral Research, 37(4), 643–648. 10.1002/jbmr.4497

Broulik, P., Urbánek, V., & Libanský, P. (2018). Eighteen-Year Effect of Androgen Therapy on Bone Mineral Density in Trans(gender) Men. Hormone and Metabolic Research, 50(02), 133–137. 10.1055/s-0043-118747

Brown, A., Montagner-Moraes, S., & Hamilton, B. (2024a). The Effects of Gender-Affirming Hormone Treatment on Transgender Musculoskeletal Health: A Protocol for a Systematic Review and Meta-Analysis. Open Science Framework. 10.17605/OSF.IO/42WY3

Brown, A., Montagner-Moraes, S., & Hamilton, B. (2024b). The effects of gender-affirming hormone treatment on transgender musculoskeletal health: a systematic review and meta-analysis. CRD42024573102. Retrieved 26th July from https://www.crd.york.ac.uk/prospero/display_record.php?ID=CRD42024573102

Brown, A., Montagner-Moraes, S., & Hamilton, B. (2025). The Effects of Gender-Affirming Hormone Treatment on Transgender Musculoskeletal Health: A Systematic Review and Meta-Analysis. Open Science Framework. 10.17605/OSF.IO/42WY3

Brown, A., Montagner-Moraes, S., Hu, K., Singh, J., Charlton, L., Barrett, J., & Hamilton, B. R. (2025). The effects of gender affirming hormone treatment on transgender women’s musculoskeletal health: a systematic review and meta-analysis. International Journal of Transgender Health, 1-28. 10.1080/26895269.2025.2590036

Chan, K. J., Jolly, D., Liang, J. J., Weinand, J. D., & Safer, J. D. (2018). Estrogen Levels Do Not Rise With Testosterone Treatment For Transgender Men. Endocrine Practice, 24(4), 329–333. 10.4158/ep-2017-0203

Cheung, A. S., Zwickl, S., Miller, K., Nolan, B. J., Wong, A. F. Q., Jones, P., & Eynon, N. (2024). The Impact of Gender-Affirming Hormone Therapy on Physical Performance. The Journal of Clinical Endocrinology & Metabolism, 109(2), e455–e465. 10.1210/clinem/dgad414

Chiccarelli, E., Aden, J., Ahrendt, D., & Smalley, J. (2023). Fit Transitioning: When Can Transgender Airmen Fitness Test in Their Affirmed Gender? Mil Med, 188(7-8), e1588–e1595. 10.1093/milmed/usac320

Cohen, J. (1968). Weighted kappa: Nominal scale agreement provision for scaled disagreement or partial credit. Psychological bulletin, 70(4), 213–220. 10.1037/h0026256

Cole, Z., Dennison, E., & Cooper, C. (2008). Update on the treatment of post-menopausal osteoporosis. Br Med Bull, 86, 129–143. 10.1093/bmb/ldn017

Coleman, E., Bockting, W., Botzer, M., Cohen-Kettenis, P., DeCuypere, G., Feldman, J., Fraser, L., Green, J., Knudson, G., & Meyer, W. J. (2012). Standards of care for the health of transsexual, transgender, and gender-nonconforming people, version 7. International Journal of Transgenderism, 13(4), 165–232. 10.1080/15532739.2011.700873

Dobrolińska, M., van der Tuuk, K., Vink, P., van den Berg, M., Schuringa, A., Monroy-Gonzalez, A. G., García, D. V., Schultz, W. C. M. W., & Slart, R. H. J. A. (2019). Bone Mineral Density in Transgender Individuals After Gonadectomy and Long-Term Gender-Affirming Hormonal Treatment. The journal of sexual medicine, 16(9), 1469–1477. 10.1016/j.jsxm.2019.06.006

Doi, S. A. R., Barendregt, J. J., Khan, S., Thalib, L., & Williams, G. M. (2015). Advances in the meta-analysis of heterogeneous clinical trials I: The inverse variance heterogeneity model. Contemporary Clinical Trials, 45, 130–138. 10.1016/j.cct.2015.05.009

Doi, S. A. R., Furuya-Kanamori, L., Thalib, L., & Barendregt, J. J. (2017). Meta-analysis in evidence-based healthcare. International Journal of Evidence-Based Healthcare, 15(4), 152–160. 10.1097/xeb.0000000000000125

Elbers, J. M., Asscheman, H., Seidell, J. C., & Gooren, L. J. (1999). Effects of sex steroid hormones on regional fat depots as assessed by magnetic resonance imaging in transsexuals. Am J Physiol, 276(2), E317–325. 10.1152/ajpendo.1999.276.2.E317

Fighera, T. M., da Silva, E., Lindenau, J. D., & Spritzer, P. M. (2018). Impact of cross-sex hormone therapy on bone mineral density and body composition in transwomen. Clin Endocrinol (Oxf), 88(6), 856–862. 10.1111/cen.13607

Fighera, T. M., Ziegelmann, P. K., Rasia Da Silva, T., & Spritzer, P. M. (2019). Bone Mass Effects of Cross-Sex Hormone Therapy in Transgender People: Updated Systematic Review and Meta-Analysis. Journal of the Endocrine Society, 3(5), 943–964. 10.1210/js.2018-00413

Follmann, D., Elliott, P., Suh, I., & Cutler, J. (1992). Variance imputation for overviews of clinical trials with continuous response. Journal of Clinical Epidemiology, 45(7), 769–773. 10.1016/0895-4356(92)90054-q

Foster Skewis, L., Bretherton, I., Leemaqz, S. Y., Zajac, J. D., & Cheung, A. S. (2021). Short-Term Effects of Gender-Affirming Hormone Therapy on Dysphoria and Quality of Life in Transgender Individuals: A Prospective Controlled Study. Frontiers in Endocrinology, 12. 10.3389/fendo.2021.717766

Furuya-Kanamori, L., Barendregt, J. J., & Doi, S. A. R. (2018). A new improved graphical and quantitative method for detecting bias in meta-analysis. International Journal of Evidence-Based Healthcare, 16(4), 195–203. 10.1097/xeb.0000000000000141

Furuya-Kanamori, L., Xu, C., Lin, L., Doan, T., Chu, H., Thalib, L., & Doi, S. A. R. (2020). P value–driven methods were underpowered to detect publication bias: analysis of Cochrane review meta-analyses. Journal of Clinical Epidemiology, 118, 86–92. 10.1016/j.jclinepi.2019.11.011

Garner, P., Hopewell, S., Chandler, J., Maclehose, H., Schünemann, H. J., Akl, E. A., Beyene, J., Chang, S., Churchill, R., Dearness, K., Guyatt, G., Lefebvre, C., Liles, B., Marshall, R., Martínez García, L., Mavergames, C., Nasser, M., Qaseem, A., Sampson, M., Soares-Weiser, K., Takwoingi, Y., Thabane, L., Trivella, M., Tugwell, P., Welsh, E., & Wilson, E. C. (2016). When and how to update systematic reviews: consensus and checklist. *bmj*, i3507. 10.1136/bmj.i3507

Gooren, L., & Bunck, M. (2004). Transsexuals and competitive sports. European Journal of Endocrinology, 151(4), 425–429. 10.1530/eje.0.1510425

Hamilton, B., Brown, A., Montagner-Moraes, S., Comeras-Chueca, C., Bush, P. G., Guppy, F. M., & Pitsiladis, Y. P. (2024). Strength, power and aerobic capacity of transgender athletes: a cross-sectional study. British Journal of Sports Medicine, 58(11), 586–597. 10.1136/bjsports-2023-108029

Hamilton, B., Brown, A., Montagner-Moraes, S., Comeras-Chueca, C., Bush, P. G., Guppy, F. M., & Pitsiladis, Y. P. (2024). Strength, power and aerobic capacity of transgender athletes: a cross-sectional study. Br J Sports Med, 58(11), 586–597. 10.1136/bjsports-2023-108029

Hamilton, B. R., Guppy, F. M., Barrett, J., Seal, L., & Pitsiladis, Y. (2021). Integrating transwomen athletes into elite competition: The case of elite archery and shooting. Eur J Sport Sci, 21(11), 1500–1509. 10.1080/17461391.2021.1938692

Hedges, L. V., & Olkin, I. (2014). Statistical methods for meta-analysis. Academic press.

Hereford, T., Kellish, A., Samora, J. B., & Reid Nichols, L. (2024). Understanding the importance of peak bone mass. Journal of the Pediatric Orthopaedic Society of North America, 7, 100031. 10.1016/j.jposna.2024.100031

Higgins, J. P. T., Thompson, S. G., Deeks, J. J., & Altman, D. G. (2003). Measuring inconsistency in meta-analyses. bmj, 327(7414), 557–560. 10.1136/bmj.327.7414.557

Iwamoto, S. J., Rice, J. D., Moreau, K. L., Cornier, M.-A., Wierman, M. E., Mancuso, M. P., Gebregzabheir, A., Hammond, D. B., & Rothman, M. S. (2024). The association of gender-affirming hormone therapy duration and body mass index on bone mineral density in gender diverse adults. Journal of Clinical & Translational Endocrinology, 36, 100348. 10.1016/j.jcte.2024.100348

Ji, M. X., & Yu, Q. (2015). Primary osteoporosis in postmenopausal women. Chronic Diseases and Translational Medicine, 1(1), 9–13. 10.1016/j.cdtm.2015.02.006

Jones, B. A., Arcelus, J., Bouman, W. P., & Haycraft, E. (2017). Sport and Transgender People: A Systematic Review of the Literature Relating to Sport Participation and Competitive Sport Policies. Sports Medicine, 47(4), 701–716. 10.1007/s40279-016-0621-y

Kasperk, C., Helmboldt, A., Börcsök, I., Heuthe, S., Cloos, O., Niethard, F., & Ziegler, R. (1997). Skeletal Site-Dependent Expression of the Androgen Receptor in Human Osteoblastic Cell Populations. Calcified Tissue International, 61(6), 464–473. 10.1007/s002239900369

Klaver, M., de Mutsert, R., Wiepjes, C. M., Twisk, J. W. R., den Heijer, M., Rotteveel, J., & Klink, D. T. (2018). Early Hormonal Treatment Affects Body Composition and Body Shape in Young Transgender Adolescents. The journal of sexual medicine, 15(2), 251–260. 10.1016/j.jsxm.2017.12.009

Kuyper, L., & Wijsen, C. (2014). Gender Identities and Gender Dysphoria in the Netherlands. Archives of Sexual Behavior, 43(2), 377–385. 10.1007/s10508-013-0140-y

Lee, E., Dobbins, M., Decorby, K., McRae, L., Tirilis, D., & Husson, H. (2012). An optimal search filter for retrieving systematic reviews and meta-analyses. BMC medical research methodology, 12(1), 51. 10.1186/1471-2288-12-51

Lu, J., Shin, Y., Yen, M.-S., & Sun, S. S. (2016). Peak Bone Mass and Patterns of Change in Total Bone Mineral Density and Bone Mineral Contents From Childhood Into Young Adulthood. Journal of Clinical Densitometry, 19(2), 180–191. 10.1016/j.jocd.2014.08.001

Lundberg, T. R., Tryfonos, A., Eriksson, L. M. J., Rundqvist, H., Rullman, E., Holmberg, M., Maqdasy, S., Linge, J., Leinhard, O. D., Arver, S., Andersson, D. P., Wiik, A., & Gustafsson, T. (2024). Longitudinal changes in regional fat and muscle composition and cardiometabolic biomarkers over 5 years of hormone therapy in transgender individuals. Journal of Internal Medicine, n/a(n/a). 10.1111/joim.20039

Lundberg, T. R., Tucker, R., McGawley, K., Williams, A. G., Millet, G. P., Sandbakk, Ø., Howatson, G., Brown, G. A., Carlson, L. A., Chantler, S., Chen, M. A., Heffernan, S. M., Heron, N., Kirk, C., Murphy, M. H., Pollock, N., Pringle, J., Richardson, A., Santos-Concejero, J., Stebbings, G. K., Christiansen, A. V., Phillips, S. M., Devine, C., Jones, C., Pike, J., & Hilton, E. N. (2024). The International Olympic Committee framework on fairness, inclusion and nondiscrimination on the basis of gender identity and sex variations does not protect fairness for female athletes. Scand J Med Sci Sports, 34(3), e14581. 10.1111/sms.14581

Maher, C. G., Sherrington, C., Herbert, R. D., Moseley, A. M., & Elkins, M. (2003). Reliability of the PEDro scale for rating quality of randomized controlled trials. Phys Ther, 83(8), 713–721. 12882612

Mohamad, N. V., Soelaiman, I.-N., & Chin, K.-Y. (2016). A concise review of testosterone and bone health. *Clinical Interventions in Aging*, Volume 11, 1317–1324. 10.2147/cia.s115472

Moher, D., Liberati, A., Tetzlaff, J., & Altman, D. G. (2009). Preferred Reporting Items for Systematic Reviews and Meta-Analyses: The PRISMA Statement. PLoS Medicine, 6(7), e1000097. 10.1371/journal.pmed.1000097

Mueller, A., Haeberle, L., Zollver, H., Claassen, T., Kronawitter, D., Oppelt, P. G., Cupisti, S., Beckmann, M. W., & Dittrich, R. (2010). Effects of Intramuscular Testosterone Undecanoate on Body Composition and Bone Mineral Density in Female-to-Male Transsexuals. The journal of sexual medicine, 7(9), 3190–3198. 10.1111/j.1743-6109.2010.01912.x

Murad, M. H., Elamin, M. B., Garcia, M. Z., Mullan, R. J., Murad, A., Erwin, P. J., & Montori, V. M. (2010). Hormonal therapy and sex reassignment: A systematic review and meta-analysis of quality of life and psychosocial outcomes. Clinical endocrinology, 72(2), 214–231. 10.1111/j.1365-2265.2009.03625.x.

Pelusi, C., Costantino, A., Martelli, V., Lambertini, M., Bazzocchi, A., Ponti, F., Battista, G., Venturoli, S., & Meriggiola, M. C. (2014). Effects of Three Different Testosterone Formulations in Female-to-Male Transsexual Persons. The journal of sexual medicine, 11(12), 3002–3011. 10.1111/jsm.12698

Pelusi, C., Costantino, A., Martelli, V., Lambertini, M., Bazzocchi, A., Ponti, F., Battista, G., Venturoli, S., & Meriggiola, M. C. (2014). Effects of three different testosterone formulations in female-to-male transsexual persons. J Sex Med, 11(12), 3002–3011. 10.1111/jsm.12698

Raggatt, L. J., & Partridge, N. C. (2010). Cellular and Molecular Mechanisms of Bone Remodeling. Journal of Biological Chemistry, 285(33), 25103–25108. 10.1074/jbc.r109.041087

Roberts, T. A., Smalley, J., & Ahrendt, D. (2021). Effect of gender affirming hormones on athletic performance in transwomen and transmen: implications for sporting organisations and legislators. British Journal of Sports Medicine, 55(11), 577–583. 55:577–583

Sarkis-Onofre, R., Catalá-López, F., Aromataris, E., & Lockwood, C. (2021). How to properly use the PRISMA Statement. Systematic Reviews, 10(1). 10.1186/s13643-021-01671-z

Scharff, M., Wiepjes, C. M., Klaver, M., Schreiner, T., T’Sjoen, G., & den Heijer, M. (2019). Change in grip strength in trans people and its association with lean body mass and bone density. Endocr Connect, 8(7), 1020–1028. 10.1530/ec-19-0196

Schünemann, H., Brożek, J., Guyatt, G., & Oxman, A. (2013). GRADE handbook for grading quality of evidence and strength of recommendations. Updated October 2013. The GRADE Working Group, 2013. Available from guidelinedevelopment. org/handbook.

Sequeira, G. M., Kidd, K., El Nokali, N. E., Rothenberger, S. D., Levine, M. D., Montano, G. T., & Rofey, D. (2019). Early Effects of Testosterone Initiation on Body Mass Index in Transmasculine Adolescents. Journal of Adolescent Health, 65(6), 818–820. 10.1016/j.jadohealth.2019.06.009

Singh-Ospina, N., Maraka, S., Rodriguez-Gutierrez, R., Davidge-Pitts, C., Nippoldt, T. B., Prokop, L. J., & Murad, M. H. (2017). Effect of Sex Steroids on the Bone Health of Transgender Individuals: A Systematic Review and Meta-Analysis. J Clin Endocrinol Metab, 102(11), 3904–3913. 10.1210/jc.2017-01642

Smart, N. A., Waldron, M., Ismail, H., Giallauria, F., Vigorito, C., Cornelissen, V., & Dieberg, G. (2015). Validation of a new tool for the assessment of study quality and reporting in exercise training studies. International Journal of Evidence-Based Healthcare, 13(1), 9–18. 10.1097/xeb.0000000000000020

Statista. (2023). Share of People Identifying as Transgender, Gender Fluid, Non-binary, or Other Ways Worldwide as of 2023, by Country.

Sterne, J. A., Hernán, M. A., Reeves, B. C., Savović, J., Berkman, N. D., Viswanathan, M., Henry, D., Altman, D. G., Ansari, M. T., Boutron, I., Carpenter, J. R., Chan, A.-W., Churchill, R., Deeks, J. J., Hróbjartsson, A., Kirkham, J., Jüni, P., Loke, Y. K., Pigott, T. D., Ramsay, C. R., Regidor, D., Rothstein, H. R., Sandhu, L., Santaguida, P. L., Schünemann, H. J., Shea, B., Shrier, I., Tugwell, P., Turner, L., Valentine, J. C., Waddington, H., Waters, E., Wells, G. A., Whiting, P. F., & Higgins, J. P. (2016). ROBINS-I: a tool for assessing risk of bias in non-randomised studies of interventions. bmj, i4919. 10.1136/bmj.i4919

Sterne, J. A. C., Savović, J., Page, M. J., Elbers, R. G., Blencowe, N. S., Boutron, I., Cates, C. J., Cheng, H.-Y., Corbett, M. S., Eldridge, S. M., Emberson, J. R., Hernán, M. A., Hopewell, S., Hróbjartsson, A., Junqueira, D. R., Jüni, P., Kirkham, J. J., Lasserson, T., Li, T., McAleenan, A., Reeves, B. C., Shepperd, S., Shrier, I., Stewart, L. A., Tilling, K., White, I. R., Whiting, P. F., & Higgins, J. P. T. (2019). RoB 2: a revised tool for assessing risk of bias in randomised trials. *bmj*, l4898. 10.1136/bmj.l4898

Stoffers, I. E., De Vries, M. C., & Hannema, S. E. (2019). Physical Changes, Laboratory Parameters, and Bone Mineral Density During Testosterone Treatment in Adolescents with Gender Dysphoria. The journal of sexual medicine, 16(9), 1459–1468. 10.1016/j.jsxm.2019.06.014

Tack, L. J. W., Craen, M., Lapauw, B., Goemaere, S., Toye, K., Kaufman, J. M., Vandewalle, S., T’Sjoen, G., Zmierczak, H. G., & Cools, M. (2018). Proandrogenic and Antiandrogenic Progestins in Transgender Youth: Differential Effects on Body Composition and Bone Metabolism. J Clin Endocrinol Metab, 103(6), 2147–2156. 10.1210/jc.2017-02316

Tominaga, Y., Kobayashi, T., Matsumoto, Y., Moriwake, T., Oshima, Y., Okumura, M., Horii, S., Sadahira, T., Katayama, S., Iwata, T., Nishimura, S., Bekku, K., Edamura, K., Sugimoto, M., Kobayashi, Y., Watanabe, M., Namba, Y., Matsumoto, Y., Nakatsuka, M., & Araki, M. (2025). Trans men can achieve adequate muscular development through low-dose testosterone therapy: A long-term study on body composition changes. Andrology, 13(2), 275–285. 10.1111/andr.13640

Turner, A., Chen, T. C., Barber, T. W., Malabanan, A. O., Holick, M. F., & Tangpricha, V. (2004). Testosterone increases bone mineral density in female-to-male transsexuals: a case series of 15 subjects. Clin Endocrinol (Oxf), 61(5), 560–566. 10.1111/j.1365-2265.2004.02125.x

Turner, A., Chen, T. C., Barber, T. W., Malabanan, A. O., Holick, M. F., & Tangpricha, V. (2004). Testosterone increases bone mineral density in female-to-male transsexuals: a case series of 15 subjects. Clinical endocrinology, 61(5), 560–566. 10.1111/j.1365-2265.2004.02125.x

Van Caenegem, E., & TLSjoen, G. (2015). Bone in trans persons. Curr Opin Endocrinol Diabetes Obes, 22(6), 459–466. 10.1097/med.0000000000000202

Van Caenegem, E., Wierckx, K., Taes, Y., Schreiner, T., Vandewalle, S., Toye, K., Lapauw, B., Kaufman, J. M., & T’Sjoen, G. (2015). Body composition, bone turnover, and bone mass in trans men during testosterone treatment: 1-year follow-up data from a prospective case–controlled study (ENIGI). European Journal of Endocrinology, 172(2), 163–171. 10.1530/eje-14-0586

van der Loos, M., Vlot, M. C., Klink, D. T., Hannema, S. E., den Heijer, M., & Wiepjes, C. M. (2023). Bone Mineral Density in Transgender Adolescents Treated With Puberty Suppression and Subsequent Gender-Affirming Hormones. JAMA Pediatr, 177(12), 1332–1341. 10.1001/jamapediatrics.2023.4588

van der Loos, M. A., Hellinga, I., Vlot, M. C., Klink, D. T., den Heijer, M., & Wiepjes, C. M. (2021). Development of Hip Bone Geometry During Gender-Affirming Hormone Therapy in Transgender Adolescents Resembles That of the Experienced Gender When Pubertal Suspension Is Started in Early Puberty. J Bone Miner Res, 36(5), 931–941. 10.1002/jbmr.4262

Van Kesteren, P., Lips, P., Deville, W., Popp-Snijders, C., Asscheman, H., Megens, J., & Gooren, L. (1996). The effect of one-year cross-sex hormonal treatment on bone metabolism and serum insulin-like growth factor-1 in transsexuals. The Journal of Clinical Endocrinology & Metabolism, 81(6), 2227–2232. 10.1210/jcem.81.6.8964856

van Kesteren, P., Lips, P., Gooren, L. J., Asscheman, H., & Megens, J. (1998). Long-term follow-up of bone mineral density and bone metabolism in transsexuals treated with cross-sex hormones. Clin Endocrinol (Oxf), 48(3), 347–354. 10.1046/j.1365-2265.1998.00396.x

Van Kesteren, P., Lips, P., Gooren, L. J. G., Asscheman, H., & Megens, J. (1998). Long-term follow-up of bone mineral density and bone metabolism in transsexuals treated with cross-sex hormones. Clinical endocrinology, 48(3), 347–354. 10.1046/j.1365-2265.1998.00396.x

VanderWeele, T. J., & Ding, P. (2017). Sensitivity analysis in observational research: introducing the E-value. Annals of internal medicine, 167(4), 268–274.

Vlot, M. C., Klink, D. T., den Heijer, M., Blankenstein, M. A., Rotteveel, J., & Heijboer, A. C. (2017). Effect of pubertal suppression and cross-sex hormone therapy on bone turnover markers and bone mineral apparent density (BMAD) in transgender adolescents. Bone, 95, 11–19. 10.1016/j.bone.2016.11.008

Vlot, M. C., Wiepjes, C. M., Jongh, R. T., T’Sjoen, G., Heijboer, A. C., & den Heijer, M. (2019). Gender-Affirming Hormone Treatment Decreases Bone Turnover in Transwomen and Older Transmen [Article]. Journal of Bone & Mineral Research, 34(10), 1862–1872. 10.1002/jbmr.3762

White Hughto, J. M., & Reisner, S. L. (2016). A Systematic Review of the Effects of Hormone Therapy on Psychological Functioning and Quality of Life in Transgender Individuals. Transgender Health, 1(1), 21–31. 10.1089/trgh.2015.0008

Wiepjes, C. M., Blok, C. J. M., Staphorsius, A. S., Nota, N. M., Vlot, M. C., Jongh, R. T., & Heijer, M. (2020). Fracture Risk in Trans Women and Trans Men Using Long-Term Gender-Affirming Hormonal Treatment: A Nationwide Cohort Study [Article]. Journal of Bone & Mineral Research, 35(1), 64–70. 10.1002/jbmr.3862

Wiepjes, C. M., de Jongh, R. T., de Blok, C. J. M., Vlot, M. C., Lips, P., Twisk, J. W. R., & den Heijer, M. (2019). Bone Safety During the First Ten Years of Gender-Affirming Hormonal Treatment in Transwomen and Transmen [Article]. Journal of Bone & Mineral Research, 34(3), 447–454. 10.1002/jbmr.3612

Wiepjes, C. M., Vlot, M. C., Klaver, M., Nota, N. M., de Blok, C. J. M., de Jongh, R. T., Lips, P., Heijboer, A. C., Fisher, A. D., Schreiner, T., T’Sjoen, G., & den Heijer, M. (2017). Bone Mineral Density Increases in Trans Persons After 1 Year of Hormonal Treatment: A Multicenter Prospective Observational Study [Article]. Journal of Bone & Mineral Research, 32(6), 1252–1260. 10.1002/jbmr.3102

Wierckx, K., Van Caenegem, E., Schreiner, T., Haraldsen, I., Fisher, A., Toye, K., Kaufman, J. M., & T’Sjoen, G. (2014). Cross-Sex Hormone Therapy in Trans Persons Is Safe and Effective at Short-Time Follow-Up: Results from the European Network for the Investigation of Gender Incongruence. The journal of sexual medicine, 11(8), 1999–2011. 10.1111/jsm.12571

Wiik, A., Lundberg, T. R., Rullman, E., Andersson, D. P., Holmberg, M., Mandić, M., Brismar, T. B., Dahlqvist Leinhard, O., Chanpen, S., & Flanagan, J. N. (2020). Muscle strength, size, and composition following 12 months of gender-affirming treatment in transgender individuals. The Journal of Clinical Endocrinology & Metabolism, 105(3), e805–e813. 10.1210/clinem/dgz24

Wylie, K., Barrett, J., Besser, M., Bouman, W. P., Bridgman, M., Clayton, A., Green, R., Hamilton, M., Hines, M., & Ivbijaro, G. (2014). Good practice guidelines for the assessment and treatment of adults with gender dysphoria. Sexual and Relationship Therapy, 29(2), 154–214. 10.1080/14681994.2014.883353

Xu, K., Fu, Y., Cao, B., & Zhao, M. (2022). Association of Sex Hormones and Sex Hormone-Binding Globulin Levels With Bone Mineral Density in Adolescents Aged 12-19 Years. Front Endocrinol (Lausanne), 13, 891217. 10.3389/fendo.2022.891217

Yun, Y., Kim, D., & Lee, E. S. (2021). Effect of Cross-Sex Hormones on Body Composition, Bone Mineral Density, and Muscle Strength in Trans Women. J Bone Metab, 28(1), 59–66. 10.11005/jbm.2021.28.1.59

Zhang, W., Cui, Z., Shen, D., Gao, L., & Li, Q. (2025). Testosterone levels positively linked to muscle mass but not strength in adult males aged 20-59 years: a cross-sectional study. Frontiers in physiology, 16. 10.3389/fphys.2025.1512268

