## Supplemental Figures and Tables for "The Effects of Gender Affirming Hormone Treatment on Transgender Men’s Musculoskeletal Health: A Systematic Review and Meta-Analysis"

**Supplementary Table 1** Quality of Evidence Assessment

| No. of Studies | Design | RoB | Indirectness of patients, intervention, and comparator | Imprecision | Inconsistency | Publication  Bias | **Quality of Evidence** |
| --- | --- | --- | --- | --- | --- | --- | --- |
| Longitudinal | | | | | | | |
| Femoral Neck | | | | | | | |
| 7 Studies (n=1005) | Observational | Moderate^1^ | Some^2^ | None | Large^4^ | Some^6^ | **Low** |
| Lumbar Spine | | | | | | | |
| 16 Studies (n=1152) | Observational | Moderate^1^ | Some^2^ | None | None | Some^5^ | **Low** |
| Total Hip | | | | | | | |
| 11 studies (n=1110) | Observational | Moderate^1^ | Some^2^ | None | Moderate^4^ | Some^5^ | **Low** |
| BMI | | | | | | | |
| 17 Studies (n=643) | Observational | Moderate^1^ | Some^2^ | None | None^4^ | None | **Moderate** |
| Body Mass | | | | | | | |
| 13 studies (n=260) | Observational | Moderate^1^ | Some^2^ | Some^3^ | None^4^ | None | **Moderate** |
| Fat Mass | | | | | | | |
| 11 Studies (n=245) | Observational | Low^1^ | Some^2^ | None | None^4^ | None | **High** |
| Body Fat % | | | | | | | |
| 9 Studies (n=142) | Observational | Moderate^1^ | Some^2^ | Some^3^ | Low | Some^5,6^ | **Moderate** |
| Fat-Free Mass | | | | | | | |
| 12 Studies (n=253) | Observational | Low^1^ | Some^2^ | None | None^4^ | None | **High** |
| Muscle Strength | | | | | | | |
| 9 Studies (n=410) | Observational | Serious^1^ | Some^2^ | Some^3^ | Moderate | Some^5,6^ | **Low** |
| **Cross-Sectional** | | | | | | | |
| Femoral Neck | | | | | | | |
| 2 Studies (n=47) | Observational | Moderate^1^ | Some^2^ | Some^3^ | Moderate | Some^5,6^ | **Low** |
| Lumbar Spine | | | | | | | |
| 2 Studies (n=56) | Observational | Moderate^1^ | Some^2^ | Some^3^ | Moderate | Some^5,6^ | **Low** |
| BMI | | | | | | | |
| 2 Studies (n=56) | Observational | Critical^1^ | Some^2^ | Some^3^ | Large | Some^5,6^ | **Low** |
| ^1^ Assessed via the RoBinS II tool.  ^2^ The authors conclude that the differing regimens of GAHT will affect comparators due to differing standards of care across studies.  ^3^ The authors consider there to be some imprecision due to the large confidence intervals in effect size  ^4^ The authors consider the inconsistency of results to be none due to the *I^2^* value being 0 for the outcome.  outcome.  ^5^ Funnell Plot Analysis showed the presence of one or more study outliers.  ^6^The number of studies for this outcome was below the recommended 10 studies to test for publication bias through the funnel plot method (Sterne et al., 2011) | | | | | | | |

**Supplementary Table 2:** Influence Analysis for Body Mass

| **Study** | **Group Excluded** | **Mean (95% CI)** | **Z (p)** | **Q (p)** | **I^2^ (%)** | **LFK Index** |
| --- | --- | --- | --- | --- | --- | --- |
| All | None | 0.18 (0.01, 0.34) | 2.07 (0.04) | 3.89 (0.99) | 0 | -0.09 (None) |
| Van Caenegem et al | Transgender Men | 0.14 (-0.04, 0.32) | 1.67 (0.04) | 2.81 (0.99) | 0 | -0.22 (None) |

Unless noted otherwise, all outcomes are reported as standardized effect size (g); ES, effect size; #, number; participants (#), number of exercise and control participants nested within ES's and studies; Z(p), z-score and alpha value; Q(p), Cochran's Q statistic and alpha value; I^2^ (%), I-squared.

⁎ Statistically significant (p<0.05).

**Supplementary Table 3:** Influence Analysis for Body Fat

| **Study** | **Group Excluded** | **Mean (95% CI)** | **Z (p)** | **Q (p)** | **I^2^ (%)** | **LFK Index** |
| --- | --- | --- | --- | --- | --- | --- |
| All | None | -0.21 (-0.48, 0.07) | -1.48 (0.14) | 11.63 (0.17) | 31 | -1.06 (Minor) |
| Tominaga et al | Transgender Men | -0.39 (-0.64, -0.14) | -2.80 (0.14) | 3.71 (0.81) | 0 | 1.04 (Minor) |

Unless noted otherwise, all outcomes are reported as standardized effect size (g); ES, effect size; #, number; participants (#), number of exercise and control participants nested within ES's and studies; Z(p), z-score and alpha value; Q(p), Cochran's Q statistic and alpha value; I^2^ (%), I-squared.⁎ Statistically significant (p<0.05).

**Supplementary Table 6:** Influence Analysis for Cross-Sectional Comparisons of Femoral Neck.

| **Study** | **Group Excluded** | **Mean (95% CI)** | **Z (p)** | **Q (p)** | **I^2^ (%)** | **LFK Index** |
| --- | --- | --- | --- | --- | --- | --- |
| All | None | 0.71 (-0.01, 1.44) | 0.92 (0.05) | **8.27 (0.02)*** | 75 | 0.24 (none) |
| Scharff and Wiepjes | Transgender Women | **0.99 (0.22, 1.77)** | **2.69 (0.05)** | 2.96 (0.09) | 66 | - |

Unless noted otherwise, all outcomes are reported as standardized effect size (g); ES, effect size; #, number; participants (#), number of exercise and control participants nested within ES's and studies; Z(p), z-score and alpha value; Q(p), Cochran's Q statistic and alpha value; I^2^ (%), I-squared.

⁎ Statistically significant (p<0.05).
